# Reevaluating the Role of Education in Cognitive Decline and Brain Aging: Insights from Large-Scale Longitudinal Cohorts across 33 Countries

**DOI:** 10.1101/2025.01.29.25321305

**Authors:** Anders M. Fjell, Ole Røgeberg, Øystein Sørensen, Inge K. Amlien, David Bartrés-Faz, Andreas M. Brandmaier, Gabriele Cattaneo, Sandra Düzel, Håkon Grydeland, Richard N. Henson, Simone Kühn, Ulman Lindenberger, Torkild Hovde Lyngstad, Athanasia M Mowinckel, Lars Nyberg, Alvaro Pascual-Leone, Cristina Solé-Padullés, Markus H. Sneve, Javier Solana, Marie Strømstad, Leiv Otto Watne, the Alzheimer’s Disease Neuroimaging Initiative, the Vietnam Era Twin Study of Aging, Kristine B. Walhovd, Didac Vidal-Piñeiro

## Abstract

Why education is linked to higher cognitive function in aging is fiercely debated. Leading theories propose that education reduces brain decline in aging, enhances tolerance to brain pathology, or that it does not affect cognitive decline but rather reflects higher early-life cognitive function. To test these theories, we analyzed 407.356 episodic memory scores from 170.795 participants >50 years, alongside 15.157 brain MRIs from 6.472 participants across 33 Western countries. More education was associated with better memory, larger intracranial volume and slightly larger volume of memory-sensitive brain regions. However, education did not protect against age-related decline or weakened effects of brain decline on cognition. The most parsimonious explanation for the results is that the associations reflect factors present early in life, including propensity of individuals with certain traits to pursue more education. While education has numerous benefits, the notion that it provides protection against cognitive or brain decline is not supported.

## Introduction

While the total number of people with dementia will increase massively due to population growth and ageing ^1^, the incidence seems to be declining^2,3^, and older adults have better cognitive function today than 20 years ago ^4^. One hypothesis is that this reflects broad societal and individual lifestyle changes, and that dementia incidence can be further reduced by promoting these ^1,5^. Education has repeatedly been suggested to be one such potential protective factor ^6,7^, in line with observations of robust associations between education and higher cognitive function in aging, as well as declines in dementia incidence with increasing population educational attainment ^8,9^. However, results so far are heterogeneous and point in different directions, and the specific mechanisms that could explain such a causal link are widely debated ^10^. We therefore suggest addressing these questions by conducting a large mega-analysis of longitudinal brain and cognitive studies covering a wider geographical distribution of samples.

Education could result in better cognition in aging by contributing to a lower rate of age-normative brain decline ^11^. Indeed, higher *brain maintenance* has been associated with better episodic memory^12^, and studies have found less brain pathology in older adults with higher education^13^, less brain decline in presymptomatic dementia ^14^, and less accumulation of cerebrovascular lesions ^15^. However, a recent longitudinal study investigating two independent samples did not find different rates of change in hippocampus and age-sensitive regions of the cerebral cortex in more educated participants ^16^. Alternatively, education could make people more resilient to underlying brain pathology by higher *cognitive reserve* ^17^. According to this theory, education leads to more efficient processing of cognitive tasks which in turn allows for higher performance despite age-normative levels of brain decline ^18^. Although a popular theory ^5,19^, a longitudinal study found that education did not weaken the link between hippocampal atrophy and memory change ^20^. Both the maintenance and the reserve accounts of education imply that education causally influences late-life cognition by reducing or postponing age-related decline. This is controversial, however, because even though education is associated with better cognitive function among older adults, it is not clear that more educated persons show less cognitive decline when measured longitudinally ^21,22^.

An alternative perspective holds that the association between education and cognitive performance is persistent across the adult lifespan. This contrasts with the more aging-centered views presented above. Under this alternative view, if education has a positive causal effect on cognition in aging, it would be by permanently boosting cognitive function earlier in life, causing persistent differences between educational groups. Increased compulsory schooling has been shown to elevate scores on tests of memory ^23–25^, intelligence ^26,27^ and general cognition ^28^, with effects detectable decades later ^29^. This perspective could also be consistent with a lack of causal effects of education on cognitive function, however, as those with higher initial cognitive functioning would be expected to reach higher levels of education than their peers. Hence, the topic of the role of education in cognitive function and brain health in aging is riddled with controversies ^30^.

Nonetheless, contrasting predictions can be derived from the different theories. If education improves memory in older age by shaping brain aging, we expect better preservation of memory-sensitive brain regions among individuals with higher education. If education improves cognitive reserve, we expect more tolerance to brain pathology, indexed by a lower correlation between brain decline and cognitive decline. In contrast, if the education-memory-brain relationship reflects stable individual differences, education should not correlate with either memory or brain decline. In that case, we also would expect to see selection effects, in the sense that participants with specific traits, especially higher cognitive function, are more likely to pursue further education. It is also relevant to examine whether retest effect – the tendency for performance to increase as a function of previous tests taken – is exaggerated with higher education. If more education yields cognitive reserve, this may manifest as a greater ability to take advantage of previous testing experience and to develop more efficient test taking strategies.

A major challenge in addressing these questions is that we need large, representative and heterogeneous longitudinal samples with sufficient statistical power. The geographic coverage is critical, because associations between brain, cognition, and education will vary both across time ^31^ and societies ^32–34^. For example, the population attributable fraction (PAF) of dementia due to low education was reported to vary from 1.7% in Argentina to 10.8% in Bolivia in a study comparing seven Latin American countries ^35^. To alleviate this concern, we here compiled data from several large studies, including a total of 407.356 memory tests from 170.795 participants across 33 countries across Europe, US and Israel, with up to seven follow-up sessions per person (see Figure 1). Although we do not have sufficient statistical power to systematically investigate effects of time, geography and societal differences, our approach ensures that the results are not confined to one specific time and place. Still, it is important to keep in mind that all samples come from WEIRD (Western, Educated, Industrialized, Rich, Democratic) countries, which limits generalizing conclusions to other societies. We focus on episodic memory because it is particularly sensitive to normal aging and neurodegenerative disease ^36^. To address brain mechanisms, we further analyzed 15,157 brain MRIs and concurrent memory tests from 6.472 participants across seven countries. The primary data sources were the population-based, multinational SHARE (Survey of Health, Ageing and Retirement in Europe) (https://share-eric.eu/) ^37^ and the Lifebrain consortium ^38^ (https://www.lifebrain.uio.no/), enriched with several legacy databases. SHARE uses probability sampling to obtain sample representativity, using the best available sample frame resources in each country to achieve full probability sampling, including access to population registers for most. Although geographically spread, MRI populations will vary in representativity, and hence we chose as strategy to validate the memory-results from SHARE in the MRI samples before conducting the brain analyses.

**Figure 1.**
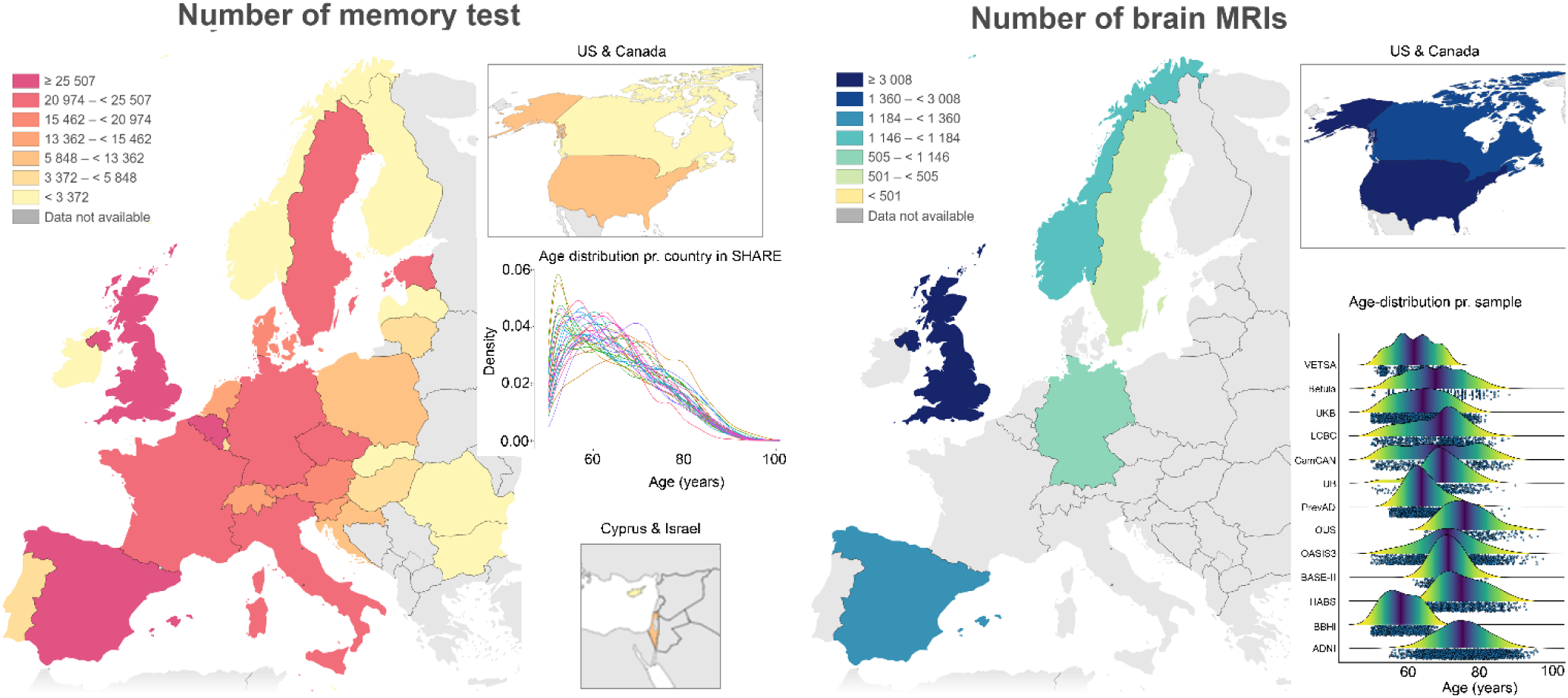
**Geographical and age distribution of samples** Left panel: Number of completed memory test sessions included across country across SHARE, Lifebrain and the other legacy datasets. The density plot shows sample age-distribution in SHARE. Right panel: Number of completed brain MRIs across countries. The plot shows the age-distribution for each dataset included. Note that the visual presentations of USA& Canada and Cyprus & Israel are not size-wise correct compared to the European map.

## Results

### SHARE cohort results

Memory was assessed with a 10-word verbal recall test, with two conditions (immediate and 5 minutes recall), using multiple versions across waves and participants ^39^. Each condition was separately included in the statistical models, yielding two observations per time point per participant. Generalized linear models with a binomial link were run using memory score as dependent variable, with the interaction between education and time since baseline as the critical term, using test type (immediate or 5-minute delay), a monotonic function of the number of previous tests taken (to control for retest effects), education, sex, country, baseline age, time since baseline, and the age × time interaction as covariates (see Online Methods for the exact model specifications). Individual-specific intercepts per participant were nested within country. Z-transformed values for age and time were used in the model fitting and converted back to natural units when showing the results. A smooth function for age allowed non-linear memory trajectories. The main outputs were the odds ratios (OR) of remembering a word compared to a reference group.

Memory scores were lower with higher baseline age, showing slightly accelerating trajectories (smoothing parameter for the combined sample = 45.8, CI: 20.7-81.5). Figure 2 (top left panel) revealed a perfect ordering of scores according to education level, with more education associated with higher scores across age. Compared to the education level used as reference (“upper secondary”), “no education” yielded OR = 0.54 compared to 1.55 for the highest category (“tertiary second stage”, Figure 3 left panel; Table 1).

**Figure 2.**
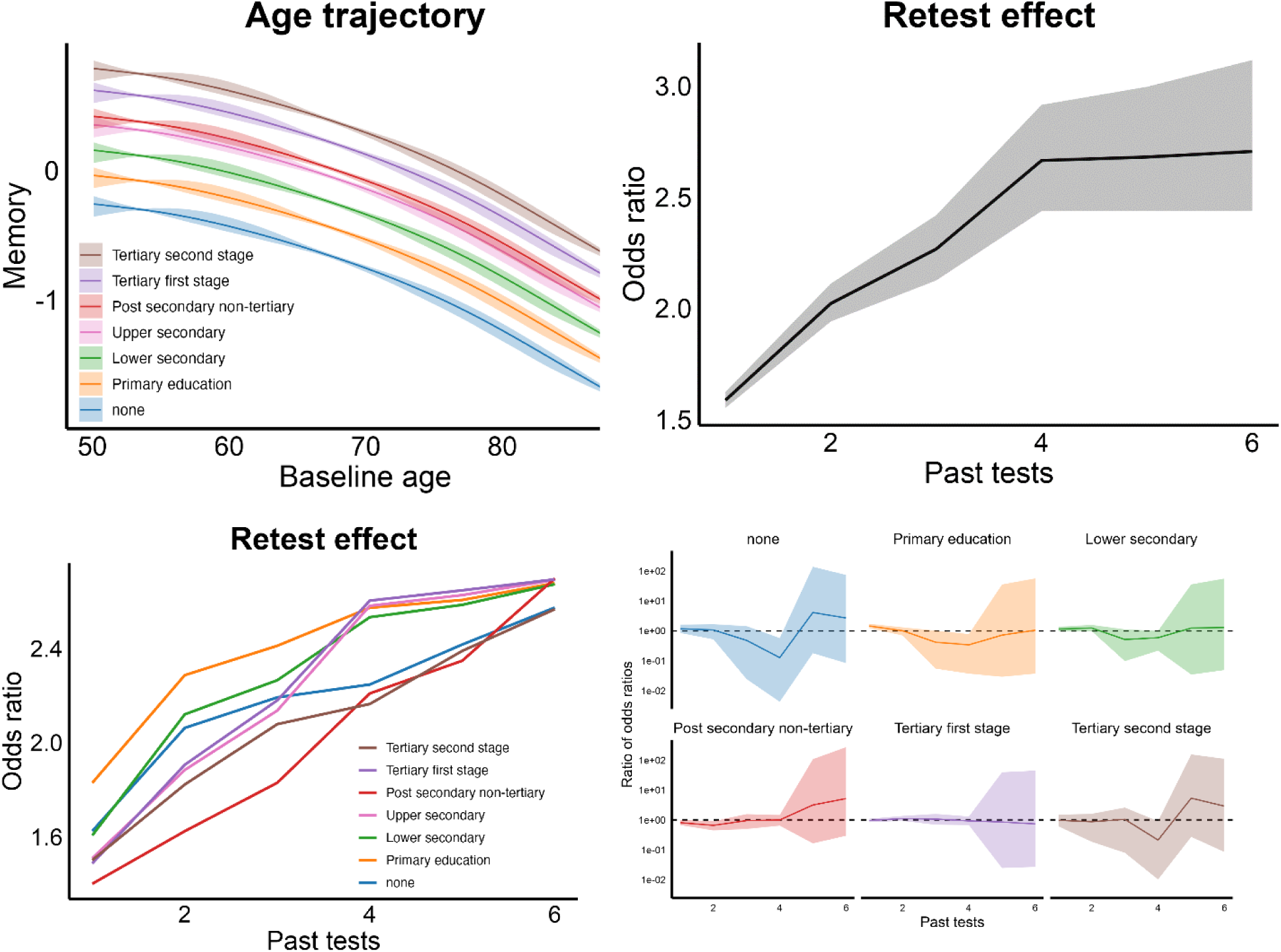
**Age, education and practice effects on memory**. Top left: Memory score trajectory as a function of baseline age. The y-axis is on the logit scale, illustrating how the linear predictor changes with varying baseline age for each education category. The legend is organized from the highest (“tertiary 2^nd^ stage”) to the lowest (“none”) level of educational attainment. Top right: Retest effects, expressed as odds ratio (y-axis) with first test session as reference and number of previous tests at the x-axis. Bottom left: Retest effects plotted for each education group. Bottom right: Comparing retest effects for each education group to the reference group by calculating Odds ratio for the given education / Odds ratio for “Upper Secondary” illustrated by the dotted horizontal line. Shaded areas denote 95% CI.

**Figure 3.**
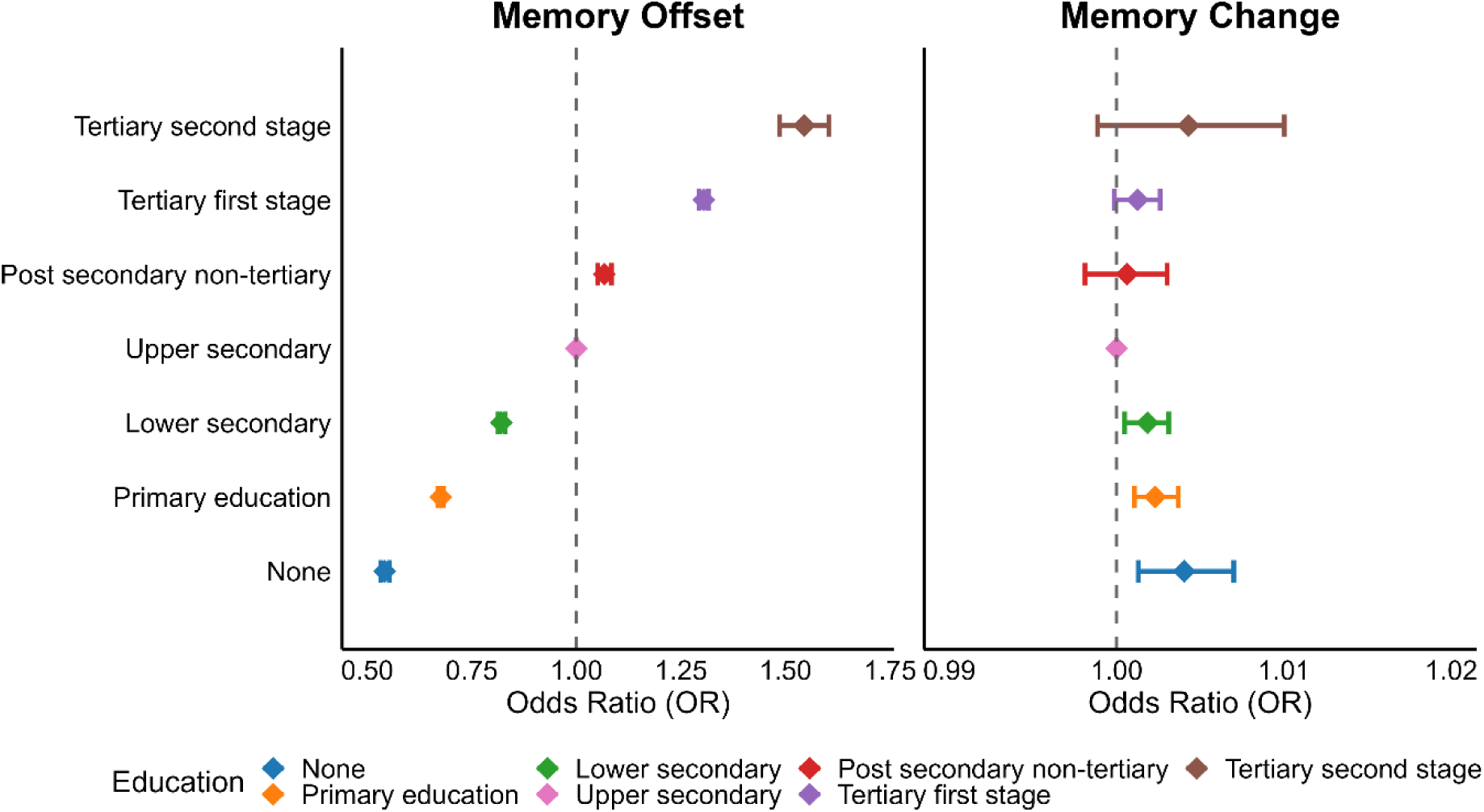
**Associations between education, memory score and memory score decline**. Left: Associations between education and memory offset scores. Right: Associations between education and decline in memory scores. ”Upper secondary” education (pink color) is used as reference, illustrated with the dashed line. Note that all memory scores are corrected for retest effects. Error bars denote 95% CI.

**Table 1.**
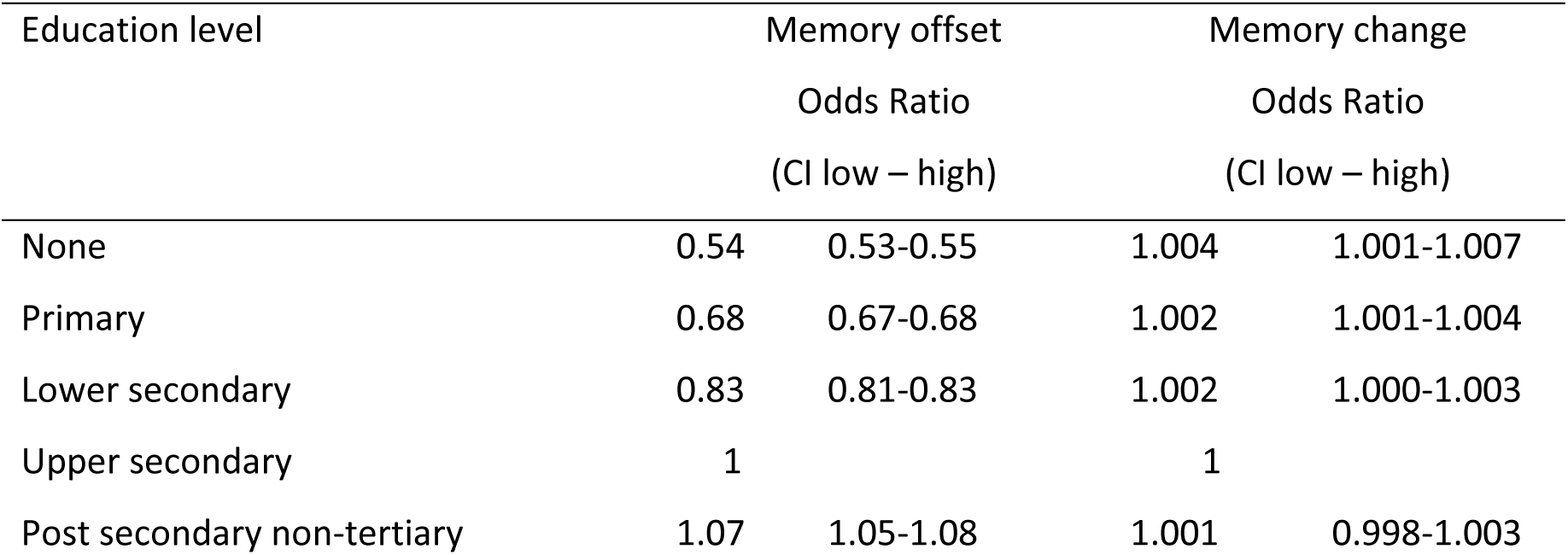
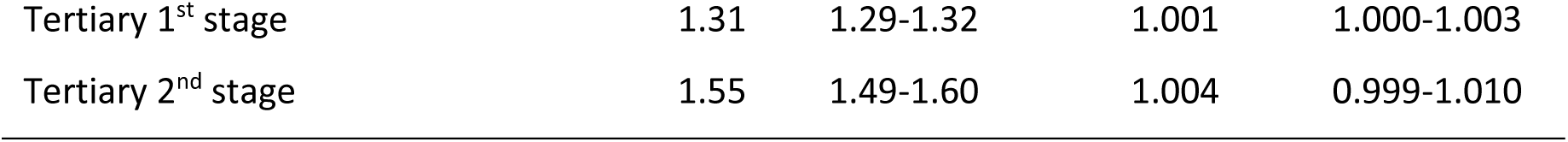
Associations between education, memory score and memory score decline. Upper secondary education is used as reference. Note that all memory scores are corrected for retest effects. Memory change (OR per year) results are presented with three decimals to allow inspection of the very weak effects. CI is 95%

| Education level | Memory offset |  | Memory change |  |
| --- | --- | --- | --- | --- |
|  | Odds Ratio |  | Odds Ratio |  |
|  | (CI low – high) |  | (CI low – high) |  |
| None | 0.54 | 0.53-0.55 | 1.004 | 1.001-1.007 |
| Primary | 0.68 | 0.67-0.68 | 1.002 | 1.001-1.004 |
| Lower secondary | 0.83 | 0.81-0.83 | 1.002 | 1.000-1.003 |
| Upper secondary | 1 |  | 1 |  |
| Post secondary non-tertiary | 1.07 | 1.05-1.08 | 1.001 | 0.998-1.003 |
| Tertiary 1<sup>st</sup> stage | 1.31 | 1.29-1.32 | 1.001 | 1.000-1.003 |
| Tertiary 2<sup>nd</sup> stage | 1.55 | 1.49-1.60 | 1.004 | 0.999-1.010 |

Retest effects were substantial and thus essential to adjust for in analyses of change. ORs increased almost linearly, from 1.5 at the first follow-up to 2.5 at the fifth (Figure 2, top right panel). There was a small negative effect of time (one year) on memory scores (OR = 0.963, CI: 0.961-0.964), slightly increasing with age (age × time OR = 0.9981, CI: 0.9980-0.9982). We assessed whether higher education was associated with less memory decline over one year (Figure 3, right panel; Table 1). Effect sizes were negligible, with all ORs < 1.005. Further, if education is associated with the ability to benefit from previous testing experience to optimize performance, individuals with more education and cognitive reserve should be able to benefit more from repeated testing more efficiently. However, there were no systematic differences in retest effects by education (Figure 4, bottom row).

**Figure 4.**
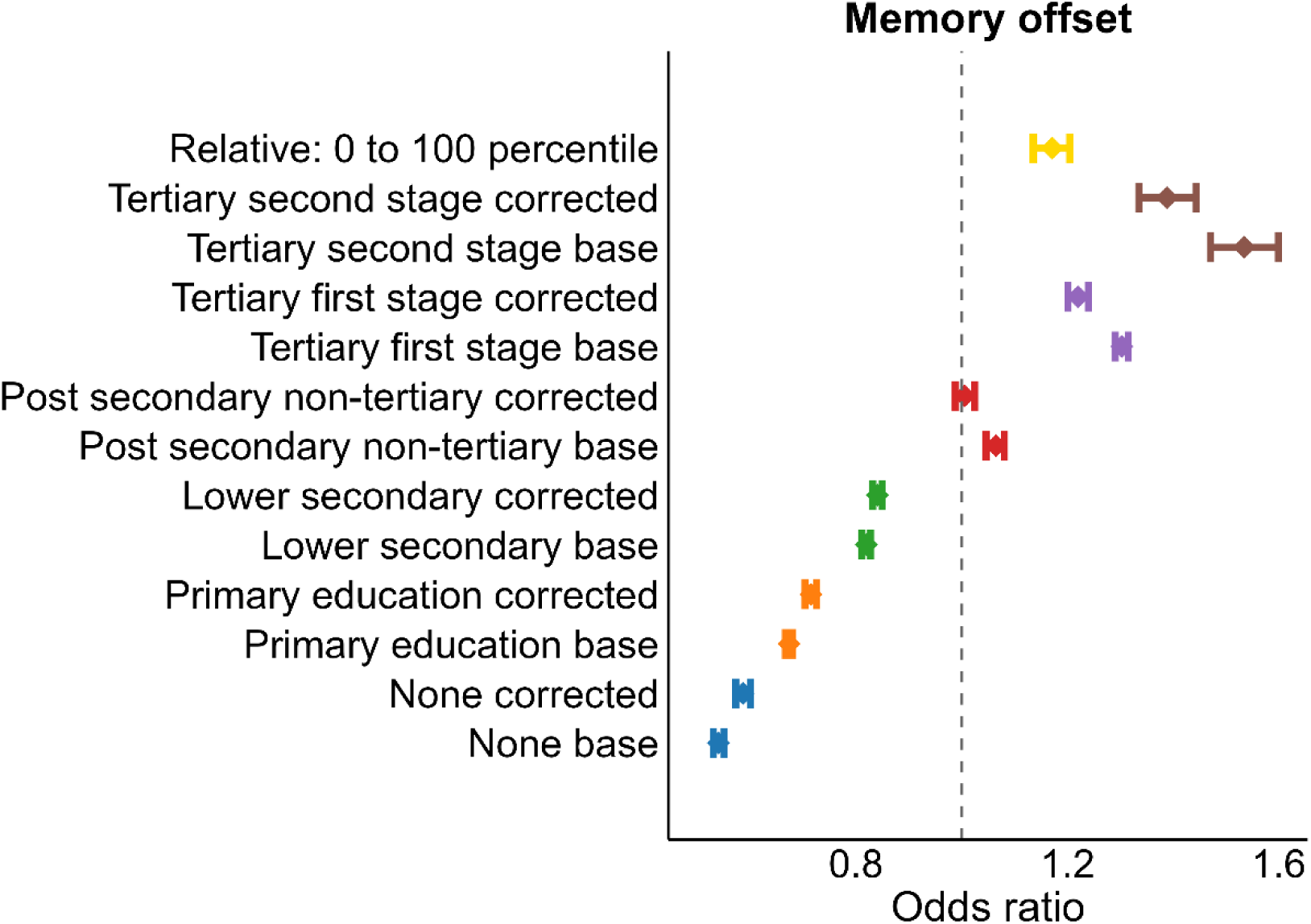
**Associations between memory, absolute and relative education**. Effects of each education category on memory compared to the reference group (“upper secondary”), illustrated with the dashed line. Models were run with (corrected) or without (base) relative education level included. Relative education was calculated as education relative to birth cohort, sex, and country, yielding a percentile score for each participant. The top row (yellow color) shows the effect of going from 0 to the 100^th^ percentile in relative education, when controlling for the influence of absolute education. Error bars denote 95% CI.

We re-ran the analyses using education relative to birth cohort in bins of a decade (1900-1909, 1910-1919, …, 1960-1969), sex, and country as measure of interest, yielding a percentile score for each participant, while controlling for absolute level of education. This provides a test of whether the education-memory associations reflect selection effects, in the sense that people are selected into education based on some unmeasured trait, that act as a common cause, and is correlated with late-life memory scores, and partially accounts for these selection effects varying between men and women from different birth cohorts in countries with widely varying educational opportunities and experiences. As seen in Figure 4, including relative education in the model reduced the associations between absolute education and memory somewhat, while relative education showed an independent, positive association with memory. The effect of going from the lowest (0) to the highest (100) percentile was associated with an OR of 1.17 (CI: 1.14-1.20) compared to the reference group (“upper secondary”).

### Brain MRI cohort results

For the brain analyses, we included 13 datasets with longitudinal MRI, memory assessments, and information about education, from seven countries across North to South of Europe, US and Canada (see Figure 2). In addition to cohort-specific inclusion and exclusion criteria, participants >50 years without cognitive impairment, neurological or psychiatric disorders were included. The initial dataset included participants with 1 to 14 MRI acquisitions with follow-up intervals spanning up to 15.8 years, and memory assessments ranging from 1 to 24 observations per participant with follow-up intervals up to 28 years. Sample characteristics are presented in Table 2, and cohort specific descriptions in Online Methods.

**Table 2.**
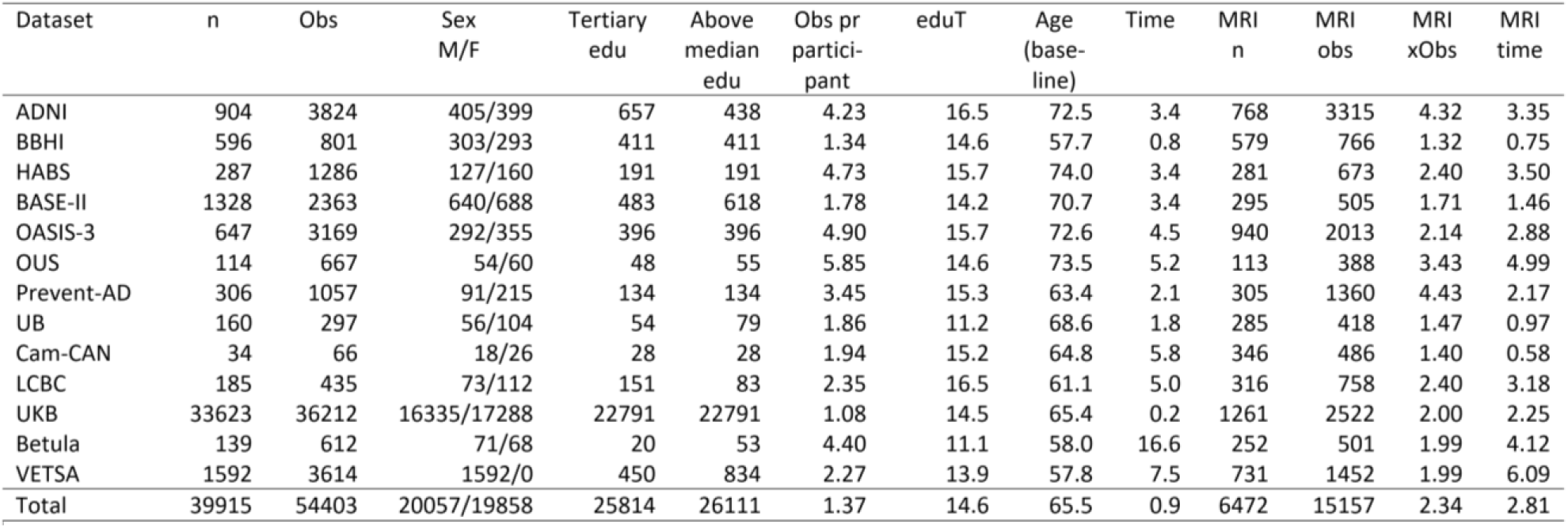
Sample characteristics for samples with MRI. . N: Number of unique participants. Obs: Total number of observations. Sex: M – Males/ F – females. Tertiary edu: Number of participants with tertiary or higher education. Above median edu: Number of participants with above median education. xObs: Obs per participant: Average number of test sessions per participant. eduT: Years of education. Time: Average maximum time in years from baseline to last follow-up. MRI: information for participants with available MRI only.

| Dataset | n | Obs | Sex<br>M/F | Tertiary<br>edu | Above<br>median<br>edu | Obs pr<br>partici-<br>pant | eduT | Age<br>(base-<br>line) | Time | MRI<br>n | MRI<br>obs | MRI<br>xObs | MRI<br>time |
| --- | --- | --- | --- | --- | --- | --- | --- | --- | --- | --- | --- | --- | --- |
| ADNI | 904 | 3824 | 405/399 | 657 | 438 | 4.23 | 16.5 | 72.5 | 3.4 | 768 | 3315 | 4.32 | 3.35 |
| BBHI | 596 | 801 | 303/293 | 411 | 411 | 1.34 | 14.6 | 57.7 | 0.8 | 579 | 766 | 1.32 | 0.75 |
| HABS | 287 | 1286 | 127/160 | 191 | 191 | 4.73 | 15.7 | 74.0 | 3.4 | 281 | 673 | 2.40 | 3.50 |
| BASE-II | 1328 | 2363 | 640/688 | 483 | 618 | 1.78 | 14.2 | 70.7 | 3.4 | 295 | 505 | 1.71 | 1.46 |
| OASIS-3 | 647 | 3169 | 292/355 | 396 | 396 | 4.90 | 15.7 | 72.6 | 4.5 | 940 | 2013 | 2.14 | 2.88 |
| OUS | 114 | 667 | 54/60 | 48 | 55 | 5.85 | 14.6 | 73.5 | 5.2 | 113 | 388 | 3.43 | 4.99 |
| Prevent-AD | 306 | 1057 | 91/215 | 134 | 134 | 3.45 | 15.3 | 63.4 | 2.1 | 305 | 1360 | 4.43 | 2.17 |
| UB | 160 | 297 | 56/104 | 54 | 79 | 1.86 | 11.2 | 68.6 | 1.8 | 285 | 418 | 1.47 | 0.97 |
| Cam-CAN | 34 | 66 | 18/26 | 28 | 28 | 1.94 | 15.2 | 64.8 | 5.8 | 346 | 486 | 1.40 | 0.58 |
| LCBC | 185 | 435 | 73/112 | 151 | 83 | 2.35 | 16.5 | 61.1 | 5.0 | 316 | 758 | 2.40 | 3.18 |
| UKB | 33623 | 36212 | 16335/17288 | 22791 | 22791 | 1.08 | 14.5 | 65.4 | 0.2 | 1261 | 2522 | 2.00 | 2.25 |
| Betula | 139 | 612 | 71/68 | 20 | 53 | 4.40 | 11.1 | 58.0 | 16.6 | 252 | 501 | 1.99 | 4.12 |
| VETSA | 1592 | 3614 | 1592/0 | 450 | 834 | 2.27 | 13.9 | 57.8 | 7.5 | 731 | 1452 | 1.99 | 6.09 |
| Total | 39915 | 54403 | 20057/19858 | 25814 | 26111 | 1.37 | 14.6 | 65.5 | 0.9 | 6472 | 15157 | 2.34 | 2.81 |

First, we tested whether the main cognitive results from SHARE replicated in the MRI cohorts. As education coding varied across samples, we could not use the same coding scheme as in SHARE, and education was hence dichotomized based on the median split for each sample, with post hoc analyses using tertiary vs. non-tertiary education as category (see *Replication analyses*). A generalized additive mixed model (*GAMM*) ^40^ was run using memory (z-normalized based on the first observation per each dataset) as dependent variable, with education, time since baseline, sex, a dummy for retest effects as fixed effects, and baseline age as smooth term. Random intercepts were included per participant and dataset while random slopes of retest and time were included for each dataset. To test memory change, an education × time interaction term was added to the model. Exact p-values are provided down to p < .001.

Like the SHARE results, while high education was associated with better memory scores (β = 0.33, SE = 0.009, p < .001), the education groups showed close to parallel changes over time (Figure 5, panels D & E). Predicted change over 10 years was z = −0.20 for high education, compared to z = −0.26 for low education (effect of education group on memory z-score change/ year: β = 0.006, SE = 0.003, p = 0.029) (for complete results, see SI). The analysis was repeated using the alternative categorization of education (tertiary vs. non-tertiary), yielding similar results.

**Figure 5.**
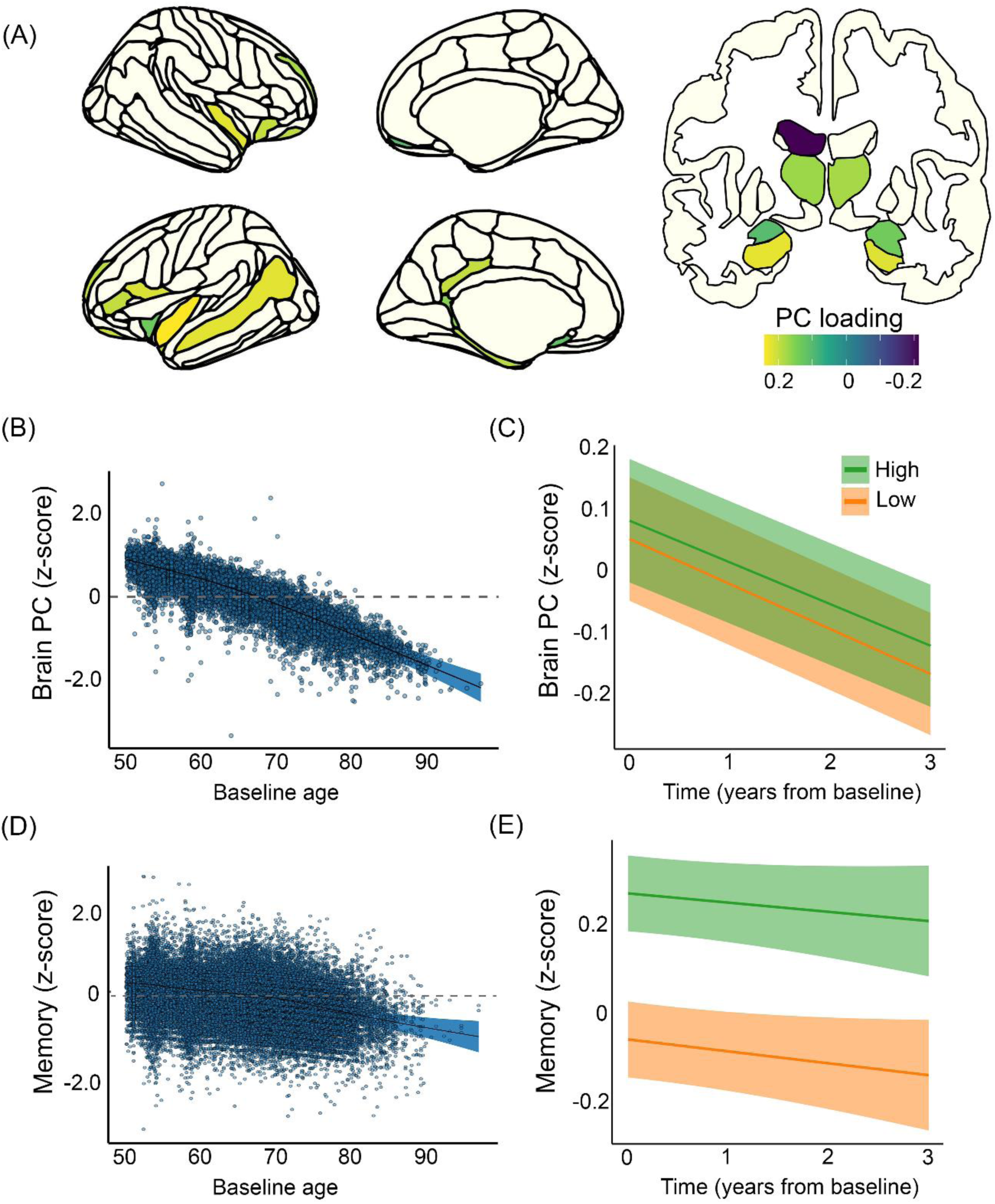
Education, brain measures and episodic memory. A: Regions where brain changes and memory changes are related (FDR < .05) are color coded by loadings on the principal component (“brain PC”). Nucleus Accumbens and left inferior lateral ventricle are not shown. B: Age-plot of the memory-sensitive PC (residuals) after accounting for sample differences. Shaded areas depict 95% CI. C: Brain change as a function of education was calculated for each education group and plotted over 3 years. Brain volumes are slightly larger for the high (green) than the low (orange) education group, but the slopes of decline are almost parallel. Shaded areas depict SE of the subject-level predictions. D: Age-plot of episodic memory (residuals) after accounting for sample differences. Shaded areas depict 95% CI. E: Episodic memory change as a function of education was calculated for each education group and plotted over 3 years. Scores are higher for the high (green) than the low (orange) education group, but the slope lines are close to parallel. SE of the subject-level predictions

We extracted a brain variable sensitive to memory change. For each participant, annual change in each of 166 brain regions was calculated and related to memory change by a series of linear mixed effects models, yielding 29 significant FDR-corrected significant regions (Figure 5, panel A). These were entered into a principal component analysis (PCA), yielding a memory-sensitive brain PC. For replication, we also used machine learning, i.e. a regularized regression model (LASSO: Least Absolute Shrinkage and Selection Operator), to predict memory based on an independent sample of 28.114 cross-sectional MRIs from UKB (*Replication analyses*).

To test the association between education and brain PC score (offset effects), a GAMM was run with education, time since baseline, sex, and estimated total intracranial volume (eTIV) as fixed effects, and baseline age and sex × baseline age as smooth terms. Random intercepts were included per participant, scanner, and dataset while random slopes of time were included for each dataset. The brain PC showed the expected negative relationship to age, slightly accelerating from about 70 years (Figure 5, panel B), and time (β = −0.07, se = 0.008, p < .001). Estimated loss in the high education group was z = −0.68 over a decade, compared to z = −0.74 for the low group (interaction effect of education × time on brain volume: β = 0.005, se = 0.002, p = .015). This means that the difference in 10-year change was z = 0.06, and the slopes were close to parallel (Figure 5, panel C). Using the alternative education categorization (tertiary vs. non-tertiary) and the LASSO-derived brain measure yielded similar results. High education was also slightly positively associated with the brain PC (β = 0.04, se = 0.02, p = .049), a relationship that was numerically stronger with the alternative education classification (β = 0.06, se = 0.02, p = .003) and weaker with the LASSO-derived brain measure (β = 0.03, se = 0.04, p = .083). When removing eTIV from the model, the estimate increased to β = 0.05 (se = 0.02, p = .019) using the median split, and to 0.07 (se = 0.02, p = .0007) using the tertiary/ non-tertiary education categorization. We tested the relationship between education and eTIV which was numerically stronger for both education classifications (median split: β = 0.12, se = .002, p < .001; tertiary/non-tertiary: β = 0.13, se = 0.02, p < .001) (Figure 6, top left).

**Figure 6.**
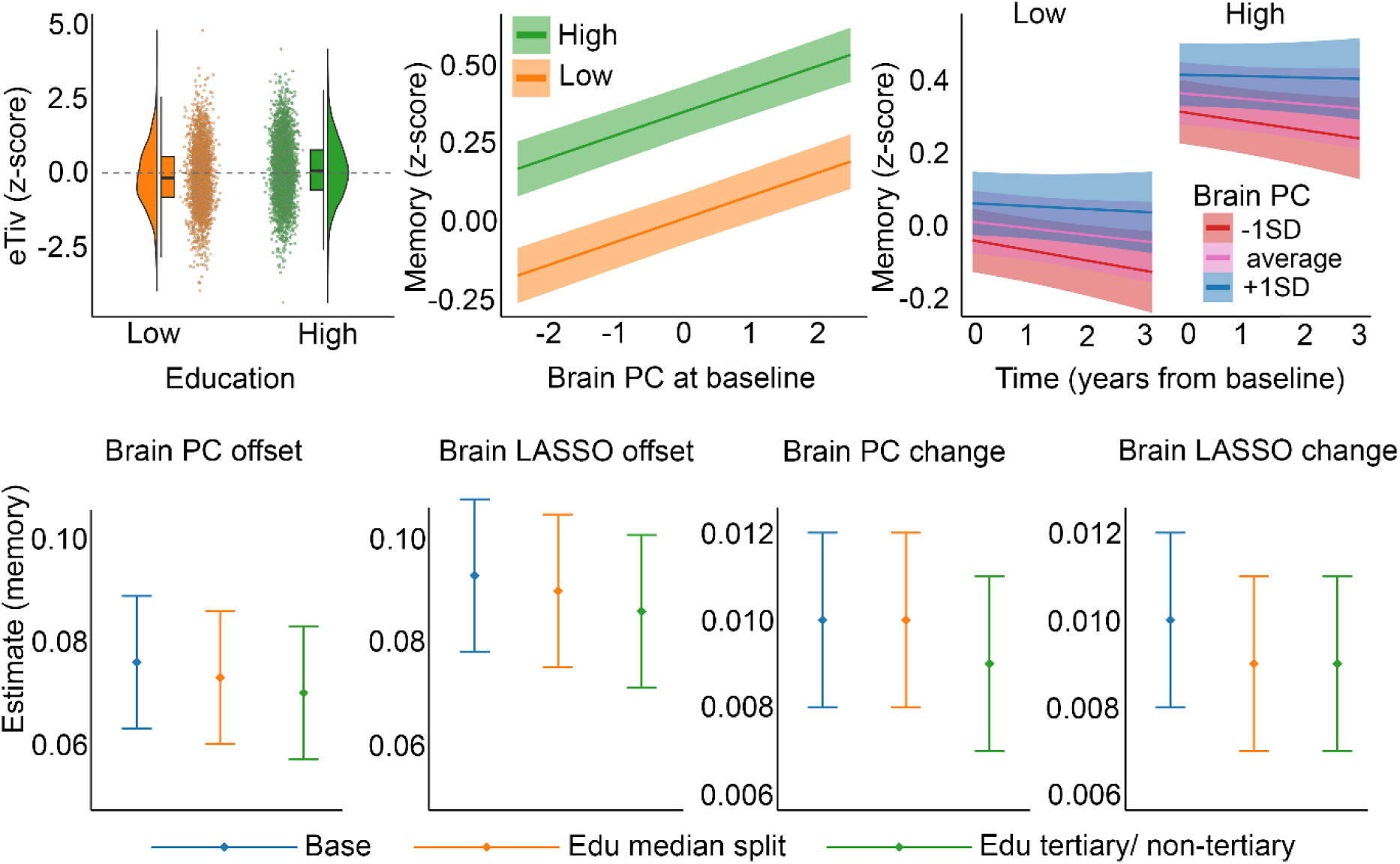
Relationships between brain, memory, and education Top row. Left: Estimated total intracranial volume (eTiv) in the high vs. low education group. Mean eTiv was significantly larger in the high education group. Middle: Relationship between brain PC at baseline and memory score separately for the high and the low education group. The brain-memory relationships are positive, but did not differ between groups. Right: Change in memory over time as a function of brain PC. More memory decline is seen for lower values of brain PC, but this relationship did not differ between education groups. Shaded areas around the lines depict SE of the subject-level predictions. **Bottom row:** Testing whether including education in the statistical models reduced the coefficients for the brain variables in predicting memory, across two brain measures and two education categorizations. Error bars depict SE. Blue: Education not included in the model. Orange/ green: Education included in the model.

Finally, we tested whether the brain-memory association varied as a function of education (see Table 3 for an overview). Higher brain PC was related to better memory (β = 0.073, se = 0.013, p < .001). As the brain PC was extracted from regions where brain change was related to memory change, the change-change relationship was given, but is still reported for completeness: β = 0.01, se = 0.002. More importantly, there were no significant education × brain PC (β = 0.01, se =0.02, p = .60) or education × brain PC × time (β = 0.004, se = 0.004, p = .43) interactions. This means that the relationship between brain and memory, and between changes in the two, did not vary as a function of education (Figure 6, top middle & right panels). The same was found using the alternative education category and the LASSO-based brain measure.

**Table 3.** Replication and control analyses. . Each of the main statistical models were run with two categorizations of education (median split, tertiary vs. non-tertiary) and two approaches to derive a brain component sensitive to memory (PCA based on memory-brain change-change relationship vs. LASSO applied to an independent dataset of cross-sectional MRIs). The main results are shown in the table, see SI for complete results. The random effect terms are not shown in the table (Random intercepts per participant and dataset, random slopes of time [and retest and for memory] for each dataset). P-values below .001 are written as ”<0.001”.

|  | Education median split |  |  |  |  |  | Education Tertiary vs. Non-tertiary |  |  |  |  |  |
| --- | --- | --- | --- | --- | --- | --- | --- | --- | --- | --- | --- | --- |
|  | Brain PCA |  |  | Brain LASSO |  |  | Brain PCA |  |  | Brain LASSO |  |  |
|  | Estimate | SE | P< | Estimate | SE | P< | Estimate | SE | P< | Estimate | SE | P< |
| **Testing brain maintenance** |  |  |  |  |  |  |  |  |  |  |  |  |
| brain ~ education category + sexFemale + s(age baseline, by = sexFemale) + time + eTIV |  |  |  |  |  |  |  |  |  |  |  |  |
| Education category | 0.038 | 0.019 | 0.049 | 0.029 | 0.035 | 0.083 | 0.060 | 0.020 | 0.003 | 0.049 | 0.018 | 0.005 |
| Time | -0.07 | 0.008 | 0.001 | -0.071 | 0.006 | 0.001 | -0.070 | 0.008 | 0.001 | -0.071 | 0.006 | 0.001 |
| Sex (female) | 0.089 | 0.024 | 0.001 | -0.445 | 0.021 | 0.001 | 0.088 | 0.024 | 0.001 | -0.446 | 0.021 | 0.001 |
| eTIV | 0.061 | 0.012 | 0.001 | -0.162 | 0.001 | 0.001 | 0.060 | 0.012 | 0.001 | -0.163 | 0.01 | 0.001 |
| brain ~ education category + sexFemale + s(age baseline, by = sexFemale) + time + eTIV + education category × time | | | | | | | | | | | | |
| Education category × time | 0.005 | 0.002 | .015 | 0.000 | 0.002 | 0.953 | 0.008 | 0.002 | 0.002 | 0.005 | 0.002 | 0.013 |
| **Testing cognitive reserve** |  |  |  |  |  |  |  |  |  |  |  |  |
| memory ~ s(age baseline, by = sexFemale) + sexFemale + brain + time + retest.dummy + eTIV |  |  |  |  |  |  |  |  |  |  |  |  |
| Brain | 0.076 | 0.013 | 0.001 | 0.093 | 0.015 | 0.001 | - | - | - | - | - | - |
| Time | -0.017 | 0.023 | 0.469 | -0.014 | 0.023 | 0.523 | - | - | - | - | - | - |
| Sex (female) | -0.385 | 0.031 | 0.001 | -0.336 | 0.031 | 0.001 | - | - | - | - | - | - |
| Retest dummy | 0.299 | 0.060 | 0.001 | 0.295 | 0.61 | 0.001 | - | - | - | - | - | - |
| eTIV | -0.006 | 0.015 | 0.66 | 0.013 | 0.015 | 0.374 | - | - | - | - | - | - |
| mem ~ s(age baseline, by = sexFemale) + sexFemale + brain + time + brain × time + retest.dummy + eTIV | | | | | | | | | | | | |
| Brain × time | 0.010 | 0.002 | 0.001 | 0.009 | 0.002 | 0.001 | - | - | - | - | - | - |
| memory ~ s(age baseline, by = sexFemale) + sexFemale + brain + time + education category + education category × brain + retest.dummy + eTIV | | | | | | | | | | | | |
| Education category × brain | 0.010 | 0.020 | 0.599 | 0.023 | 0.021 | 0.270 | -0.005 | 0.020 | 0.793 | 0.008 | 0.022 | 0.711 |
| memory ~ s(age baseline, by = sexFemale) + sexFemale + brain × time × education category + retest.dummy + eTIV | | | | | | | | | | | | |
| Brain × time × education | 0.004 | 0.004 | 0.426 | 0.004 | 0.005 | 0.392 | 0.004 | 0.004 | 0.426 | 0.008 | 0.005 | 0.063 |

### Replication analyses

The main analyses were run using the alternative categorization of education (tertiary vs. non-tertiary) and brain measure (LASSO), yielding four model specifications (Table 3). Controlling for eTIV, cross-sectional education-brain associations were relatively weak, although significant at p < .05 in three models. The education × time interaction showed small effect sizes in the same three specifications, but still significant. Effect size was largest for the PC brain measure and the tertiary/ non-tertiary categorization, with an interaction coefficient of 0.008 compared to 0.005 for the two other significant specifications. The brain × education × time interaction on memory was not significant in any specification.

As an additional set of control analyses, we tested whether the coefficients for the brain variables in predicting memory were affected by including education in the models (Figure 6, bottom panels). The coefficients changed only minimally, suggesting that the brain-memory relationships were largely independent of education (full results in SI).

## Discussion

We found that education was only minimally associated with less age-related decline in episodic memory function, not associated with any substantial reduction in the rate of age-normative structural brain decline in memory-sensitive regions, and did not increase cognitive resilience to the observed brain changes. The small magnitude of the differences in brain and memory change across educational groups contrast with the comparatively much larger differences in level. We found, in line with previous studies, that education was associated with better episodic memory scores across the age-range, slightly larger volume of the memory-sensitive brain regions, and larger intracranial volume. These associations are likely rooted in lifelong variation in brain structure and function that originate earlier in life ^30^. We also find evidence that selection effects may account for parts of the associations, in the sense that people with certain traits, such as larger brain volumes from early age as indicated by estimates of total intracranial volume and better episodic memory, tend to be selected into longer education. This selection likely varies across social and demographic dimensions as well as across features of the educational system, but it is important to note that clear patterns of associations resulted from analyses conducted on diverse samples covering a large number of WEIRD societies and age cohorts, indicating a certain degree of robustness across time and place. The implications of the results are discussed below.

### A role for education in brain and cognitive aging?

The idea that age-related cognitive decline is reduced by higher education is based on two complementary hypotheses. According to the first, education can guard against memory decline by causally influencing lifestyle factors that preserve memory-sensitive brain regions, i.e. by increased brain maintenance. While we find support for the observation that relative absence of brain decline in terms of less atrophy is linked to better episodic memory ^12^, there were, however, only minor differences in the decline trajectories of memory-sensitive brain regions across educational groups. This aligns with and extends a previous finding that educational level is not associated with differences in age-change in the brain regions most vulnerable to normal aging ^16^. In sum, these results provide a neurobiological perspective for why people with different educational attainment and different levels of memory function may still show similar rates of age-related memory decline ^21,41^ - simply put, brains change across middle- and older age in very similar ways across the entire spectrum of observed differences in education.

The second hypothesis is that education protects cognitive function through increased resilience to brain decline by building a “cognitive reserve” ^5,18,19^. This hypothesis implies that people with more education should have higher cognitive performance than expected given their observed level of brain decline ^19^. We find little support for this idea: only very small differences in the aging trajectories for memory and the memory-sensitive brain regions were observed between educational levels. Further, structural brain decline was associated with similar amounts of memory decline in more vs. less educated participants, consistent with previous research on hippocampal ^20^ and cortical ^42^ atrophy. Finally, more education was not associated with larger retest effects, which suggests that education did not come with greater ability to benefit from the specific test experience^43^. Retest effects reflect the ability to take advantage of previous testing to improve test scores. More educated participants showed greater ability to encode new information, as reflected in their higher memory scores, but this did not increase their ability to benefit from previous testing. Similar results have been found for tests of mental speed and reasoning ^44^. Taken together, the results suggest that education was not associated with less decline in brain or episodic memory in aging, and that the positive associations consequently must have been established before the age of 50 years. Although the present data do not include developmental information, we can speculate that the precursors of the differences in brain and cognition observed in aging were already present early in life, as discussed further in the next section.

### How do associations between brain volume, cognitive function and education arise?

The results revealed a robust relationship between education, higher memory function, slightly larger volumes of memory-sensitive brain regions, and larger intracranial volume. Understanding the nature of these associations is important. The most obvious explanation is that they may reflect that persons with higher cognitive abilities and larger brain volumes are more likely to select and be selected to further education ^45^. Although there were unequal opportunities and clear limitations to access to education for many of the participants in the present study ^46^, likely reducing the relationship between cognitive abilities and educational attainment, the existence of selection effects is well documented in previous studies. The present results suggest that this may account for at least a part of the relationship between education and memory function. Regardless of absolute educational attainment, participants with high education relative to other participants of the same sex, birth cohort and country of residence demonstrated better memory function decades later, consistent with the expectation that selection effects contribute to the observed relationship. Earlier-life cognition predicts cognitive function and brain health in aging ^47,48^, suggesting limited opportunities for causal effects of education beyond adolescence. Instead, selection effects driven by early-life cognitive abilities and gross aspects of brain structure may explain the life-long associations between education and cognition, also consistent with recent genetic evidence ^49,50^. Our results are also in line with a systematic review of effects of education on dementia risk, which argued that low education has a stronger association with dementia when it reflects cognitive capacity rather than privilege, and when it is associated with other risk factors across the lifespan ^51^. Furthermore, cognition-education relationships can in part be explained by neuroanatomical volume differences established in early childhood ^34^, also limiting the potential causal effects of later education. Brain structure may hence be a key phenotype along the causal pathway that leads from genetic variation to differences in cognitive function and educational attainment ^52^.

While selection effects are real, natural experiments still suggest that increased education can positively impact cognitive function ^26–28^, including memory ^23–25^. The results showed that taking selection effects into account reduced the association between education and memory only to a modest extent. Importantly, positive effects of increased education are due to early schooling, not reduced decline in aging ^29^. Our finding of similar memory-education associations across the age-range aligns with evidence that education enhances lifelong cognitive function without affecting age-related decline. Still, most cognitive intervention studies find that positive effects on cognitive scores diminish over time ^21,53^, so associations would be expected to be small when measured decades later. Thus, any early effect of education on cognition would likely need to be sustained by some mechanism that helps maintaining the initial effect, e.g. by increasing the likelihood of working in cognitively challenging occupations. According to the gravitational hypothesis, the stability of individual differences in cognition is caused by consistent exposure to the same environments over time, including social, educational, and economic contexts ^54^, see ^21,55^ for more in-depth discussions of this topic. This is in line with studies finding ‘cognitive stimulation’ at work to be associated with lower dementia risk ^56^, although this cannot explain the full association between education and less dementia ^57^. Nonetheless, individuals with higher cognitive function may pursue cognitively stimulating activities irrespective of their formal education, potentially leading to spurious associations when this is not accounted for.

An interesting aspect of the present results was the linear association between memory performance and educational attainment. If education caused cognitive scores to increase, one could expect diminishing marginal benefits with increasing duration, although this question has not been properly addressed by quasi-experimental methods ^29^. Hence, this result could reflect that selection effects are additive across the range of educational levels, but definite evidence is currently lacking. It is also interesting that this clear pattern is identified across samples covering a large number of countries and cohorts, suggesting that this entails a certain degree of robustness to societal variations across different WEIRD societies.

We observed that individuals with higher education had slightly larger volumes in memory-sensitive brain regions. Experiments have showed effects of cognitive training on both memory and relevant brain structures even in older adults ^58–60^, and it is possible that early education could lead to increased brain volumes of a magnitude similar to that observed in the present study. However, training-induced effects on brain structure are generally even more transient than those on cognition ^61,62^, making it less likely that direct effects of youth education on brain volume would persist into old age. Consistent with this, a recent study found no evidence of structural brain differences resulting from the increase in mandatory schooling from 15 to 16 years in the UK 50 years later ^63^. Instead, intracranial volume has been shown to be more strongly related to education than gray matter volume ^34^, which was also found in the current study. In fact, the association between education and intracranial volume was double the size of the association with the memory-sensitive brain component, and removing intracranial volume from the models increased the relationship between memory scores and the memory-sensitive brain PC. Since intracranial volume reaches its maximum in childhood and is unlikely to be influenced by schooling, this relationship does not reflect a causal effect of education and is a further indication that selection effects indeed play a role. Although the relationship between brain volumes and education was found to exist also independently of intracranial volume, it is most likely that the education-brain association was present early in life. Therefore, we interpret the memory-brain-education relationships observed in the present study as partly reflecting selection effects, potentially complemented by some self-reinforcing effects of early schooling.

### Considerations and future research

First, the samples cover 33 countries, and the conclusions not confined to one specific time and place. Still, we did not attempt to detect variations in associations across time ^31^ and societies ^32–35,64^, but another a multi-cohort, multi-national aging-study found relatively consistent associations between cognition and education ^65^. Second, while SHARE used probability sampling to achieve representativity, the MRI samples are generally less representative of their respective populations (e.g. ^66^). It is difficult to estimate the influence of this, but we note that the memory-education results from SHARE were replicated in the brain imaging cohorts. Further, selective attrition and mortality may affect the longitudinal estimates, although studies addressing this have largely obtained similar estimates ^21^.

Third, we used memory test scores as measures of cognition. While such scores correlate with important real-life indicators, e.g. work participation and capacity for independent living, it is not clear to what extent changes in test scores imply similar changes in daily life cognitive function (for a broader discussion, see ^67^). It cannot be ruled out that education enhanced scores by increased test-taking skills or cognitive strategies with little effect on the underlying cognitive construct. Such effects could be expected to be larger for crystallized or domain knowledge-based tests, such as vocabulary or calculus, and less for fluid tests, including list recall ^21^. Still, schooling can potentially increase fluid test performance by factors such as test-specific encoding strategies and test-taking skills, which may have little applicability to other aspects of life.

Finally, we focused on episodic memory and structural brain changes. Causal effects of education have been identified for various cognitive measures, including fluid (such as memory), crystallized (e.g. language) and compound (e.g. the *g-*factor) measures of cognition ^29^. One study found that the association between education and cognitive scores, when controlling for childhood cognition, comprised direct effects on specific cognitive skills, including memory, and was not mediated by the g-factor ^68^. Therefore, a potential extension of the current work would be to include multiple cognitive functions and examine common versus unique associations with education and brain structure. Finally, although structural brain change is predictive of memory decline in aging ^36^, other brain measures, such as those related to brain connectomics ^69^, could potentially show different relationships to education.

## Conclusion

In this large-scale, geographically spread longitudinal mega-analytic study, we find that education is robustly related to higher episodic memory function and intracranial volume, and modestly to a brain component optimized to be sensitive to memory change. However, the results do not indicate that this association is driven by slower brain aging or more resilience to structural brain change. Rather, we find evidence to suggest that the relationship is established early in life and partly is attributable to selection effects. Hence, to the extent that education may have a positive effect on episodic memory function in aging, this effect originates from earlier in life.

## Data Availability

Each dataset has different owners. Contact information to be used for requests for data access is specified in a table in the manuscript.

## Acknowledgement

The Lifebrain consortium is funded by the EU Horizon 2020 grant agreement no. 732592 (Lifebrain). The different sub-studies are supported by different sources. LCBC is supported by the European Research Council under grant agreements no. 283634 and no. 725025 (to A.M.F.) and no. 313440 (to K.B.W.), as well as the Norwegian Research Council (325878, 262453 to A.M.F.; 325001, 301395, 239889 to K.B.W.; 249931 to A.M.F & K.B.W.; 324882 to DVP; 325415 to HG), the National Association for Public Health’s dementia research program, Norway (to A.M.F.), and the University of Oslo through the UiO:Life Science convergence environment (to A.M.F). Betula is supported by a scholar grant from the Knut and Alice Wallenberg foundation to L.N. Barcelona is partially supported by a Spanish Ministry of Economy and Competitiveness grant to D.B.-F. (grant no. PSI2015-64227-R (AEI/FEDER, UE)); and to and to G.C and J.S.-S (grant no. PID-2022-139298OA-C22 (MCIN /AEI/10.13039/501100011033 / FEDER, UE)); by the Walnuts and Healthy Aging study (http://www.clinicaltrials.gov; grant no. NCT01634841) funded by the California Walnut Commission, Sacramento, California; and by an ICREA Academia 2019 award. BASE-II has been supported by the German Federal Ministry of Education and Research under grant nos 16SV5537, 16SV5837, 16SV5538, 16SV5536K, 01UW0808, 01UW0706, 01GL1716A and 01GL1716B and by the European Research Council under grant agreement no. 677804 (to S.K.). Dr. A. Pascual-Leone is partly supported by grants from the National Institutes of Health (R01AG076708), Jack Satter Foundation, and BrightFocus Foundation.

Part of the research was conducted using the UKB resource under application no. 32048. The funders had no role in study design, data collection and analysis, decision to publish or preparation of the manuscript. LOW is funded by the South-Eastern Norway Regional Health Authorities (#2017095), the Norwegian Health Association (#19536, #1513) and by Wellcome Leap’s Dynamic Resilience Program (jointly funded by Temasek Trust) (#104617).

Parts of the data used in preparation of this article were obtained from the Pre-Symptomatic Evaluation of Novel or Experimental Treatments for Alzheimer’s Disease (PREVENT-AD) program. Data were provided [in part] by OASIS-3” “OASIS-3: Principal Investigators: T. Benzinger, D. Marcus, J. Morris; NIH P50AG00561, P30NS09857781, P01AG026276, P01AG003991, R01AG043434, UL1TR000448, R01EB009352.

Parts of the data collection and sharing for this project were provided by the Cambridge Centre for Ageing and Neuroscience (CamCAN). CamCAN funding was provided by the UK Biotechnology and Biological Sciences Research Council (grant number BB/H008217/1), together with support from the UK Medical Research Council and University of Cambridge, UK.

Parts of the data are from VETSA, which is funded by the National Institute of Aging grants R01s AG018384, AG018386, AG050595, AG022381, AG076838. The content is the responsibility of the authors and does not necessarily represent official views of the NIA, NIH, or VA. U.S. Department of Veterans Affairs, Department of Defense; National Personnel Records Center, National Archives and Records Administration; Internal Revenue Service; National Opinion Research Center; National Research Council, National Academy of Sciences; and the Institute for Survey Research, Temple University provided invaluable assistance in the conduct of the VET Registry. The Cooperative Studies Program of the U.S. Department of Veterans Affairs provided financial support for development and maintenance of the Vietnam Era Twin Registry. We would also like to acknowledge the continued cooperation and participation of the members of the VET

Registry and their families

Part of the data collection and sharing was funded by the Alzheimer’s Disease Neuroimaging Initiative (ADNI) (National Institutes of Health Grant U01 AG024904) and DOD ADNI (Department of Defense award number W81XWH-12-2-0012). ADNI is funded by the National Institute on Aging, the National Institute of Biomedical Imaging and Bioengineering, and through generous contributions from the following: AbbVie, Alzheimer’s Association; Alzheimer’s Drug Discovery Foundation; Araclon Biotech; BioClinica, Inc.; Biogen; Bristol-Myers Squibb Company; CereSpir, Inc.; Cogstate; Eisai Inc.; Elan Pharmaceuticals, Inc.; Eli Lilly and Company; EuroImmun; F. Hoffmann-La Roche Ltd and its affiliated company Genentech, Inc.; Fujirebio; GE Healthcare; IXICO Ltd.; Janssen Alzheimer Immunotherapy Research & Development, LLC.; Johnson & Johnson Pharmaceutical Research & Development LLC.; Lumosity; Lundbeck; Merck & Co., Inc.; Meso Scale Diagnostics, LLC.; NeuroRx Research; Neurotrack Technologies; Novartis Pharmaceuticals Corporation; Pfizer Inc.; Piramal Imaging; Servier; Takeda Pharmaceutical Company; and Transition Therapeutics. The Canadian Institutes of Health Research is providing funds to support ADNI clinical sites in Canada. Private sector contributions are facilitated by the Foundation for the National Institutes of Health ( www.fnih.org). The grantee organization is the Northern California Institute for Research and Education, and the study is coordinated by the Alzheimer’s Therapeutic Research Institute at the University of Southern California. ADNI data are disseminated by the Laboratory for Neuro Imaging at the University of Southern California.

Parts of the data used in the preparation of this article were obtained from the Harvard Aging Brain Study (HABS - P01AG036694; https://habs.mgh.harvard.edu). The HABS study was launched in 2010, funded by the National Institute on Aging, and is led by principal investigators Reisa A. Sperling MD and Keith A. Johnson MD at Massachusetts General Hospital/Harvard Medical School in Boston, MA.

The SHARE data collection has been funded by the European Commission, DG RTD through FP5 (QLK6-CT-2001-00360), FP6 (SHARE-I3: RII-CT-2006-062193, COMPARE: CIT5-CT-2005-028857, SHARELIFE: CIT4-CT-2006-028812), FP7 (SHARE-PREP: GA N°211909, SHARE-LEAP: GA N°227822, SHARE M4: GA N°261982, DASISH: GA N°283646) and Horizon 2020 (SHARE-DEV3: GA N°676536, SHARE-COHESION: GA N°870628, SERISS: GA N°654221, SSHOC: GA N°823782, SHARE-COVID19: GA N°101015924) and by DG Employment, Social Affairs & Inclusion through VS 2015/0195, VS 2016/0135, VS 2018/0285, VS 2019/0332, VS 2020/0313, SHARE-EUCOV: GA N°101052589 and EUCOVII: GA N°101102412. Additional funding from the German Federal Ministry of Education and Research (01UW1301, 01UW1801, 01UW2202), the Max Planck Society for the Advancement of Science, the U.S. National Institute on Aging (U01_AG09740-13S2, P01_AG005842, P01_AG08291, P30_AG12815, R21_AG025169, Y1-AG-4553-01, IAG_BSR06-11, OGHA_04-064, BSR12-04, R01_AG052527-02, R01_AG056329-02, R01_AG063944, HHSN271201300071C, RAG052527A) and from various national funding sources is gratefully acknowledged (see www.share-eric.eu).

## Online Methods

### Samples

#### SHARE cohort

The Survey of Health, Ageing and Retirement in Europe is a research infrastructure for studying the effects of health, social, economic and environmental policies over the life-course of European citizens and beyond (https://share-eric.eu/) ^37^. SHARE contains observations of individuals from 50 years of age from 28 countries, recruited to be representative of the population in each country. Data for the present analyses was extracted from *easy*SHARE (release 8.0.0, February 10^th^ 2022, doi:10.6103/SHARE.easy.800), see ^70,71^ for methodological details. The *easy*SHARE release 8.8.0 is based on SHARE Waves 1, 2, 3, 4, 5, 6, 7, and 8 (DOIs:10.6103/SHARE.w1.800, 10.6103/SHARE.w2.800, 10.6103/SHARE.w3.800, 10.6103/SHARE.w4.800, 10.6103/SHARE.w5.800, 10.6103/SHARE.w6.800, 10.6103/SHARE.w7.800, 10.6103/SHARE.w8.800) ^37,72^. Participants included in the analyses participated in up to six waves of data collection. In total, we included data from 130.880 participants (mean age 64.9 years at baseline, 50.1-112.0, 59.363 males/ 71.517 females), with an average of 2.7 (SD = 1.63) waves with a mean maximum follow-up interval of 6.53 years (0.9-0-15.9, SD = 3.93). In total, 352.953 memory test sessions were included, with two test results (immediate vs. delayed recollection) for each, i.e. 705.906 memory scores went into the analyses. Respondents aged below 50 years of age (individuals recruited due to being spouses of other participants) were excluded from the sample. An overview of sample distribution as a function of timepoints, education category and age is provided in the figure below.

**Online Figure:**
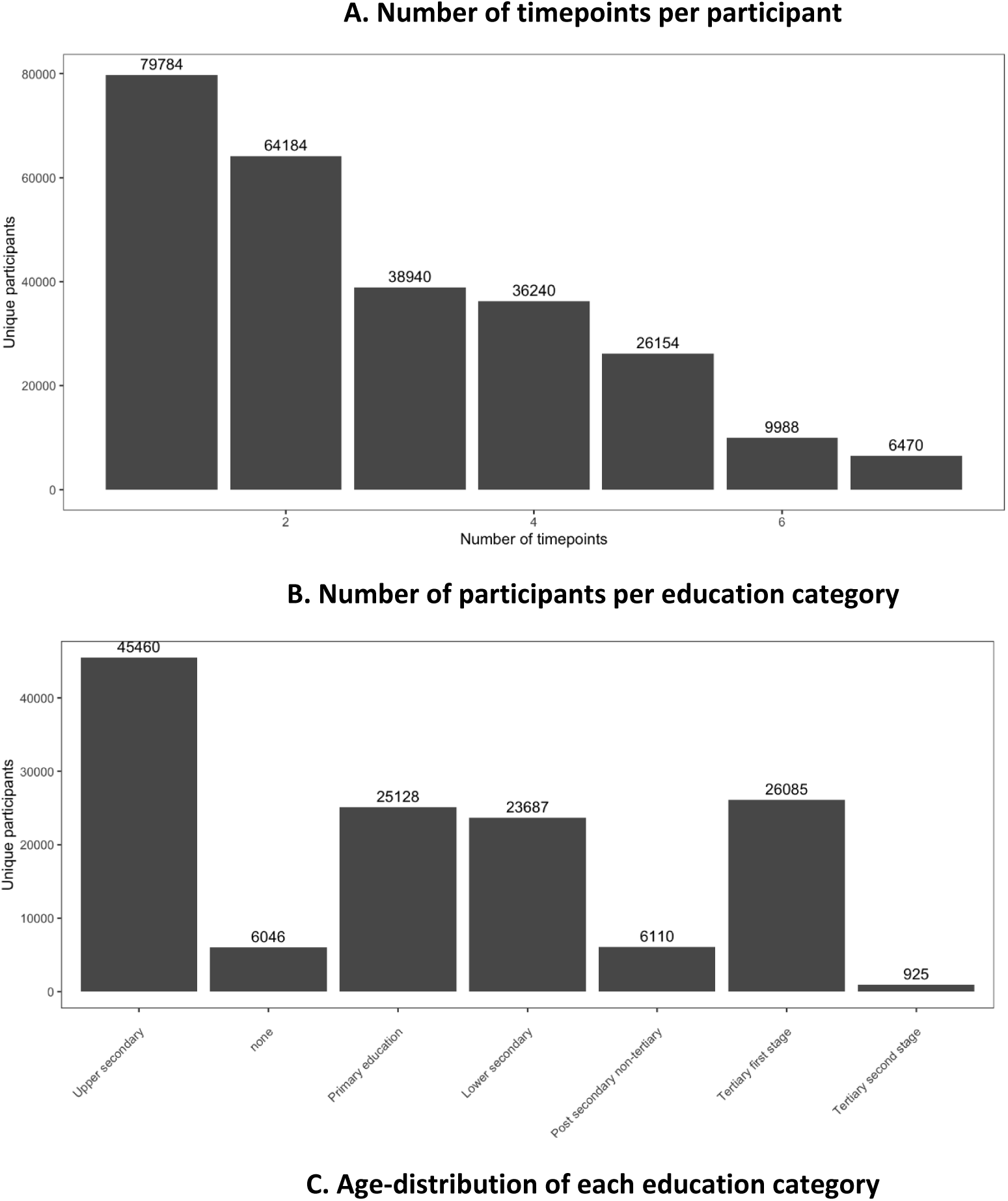
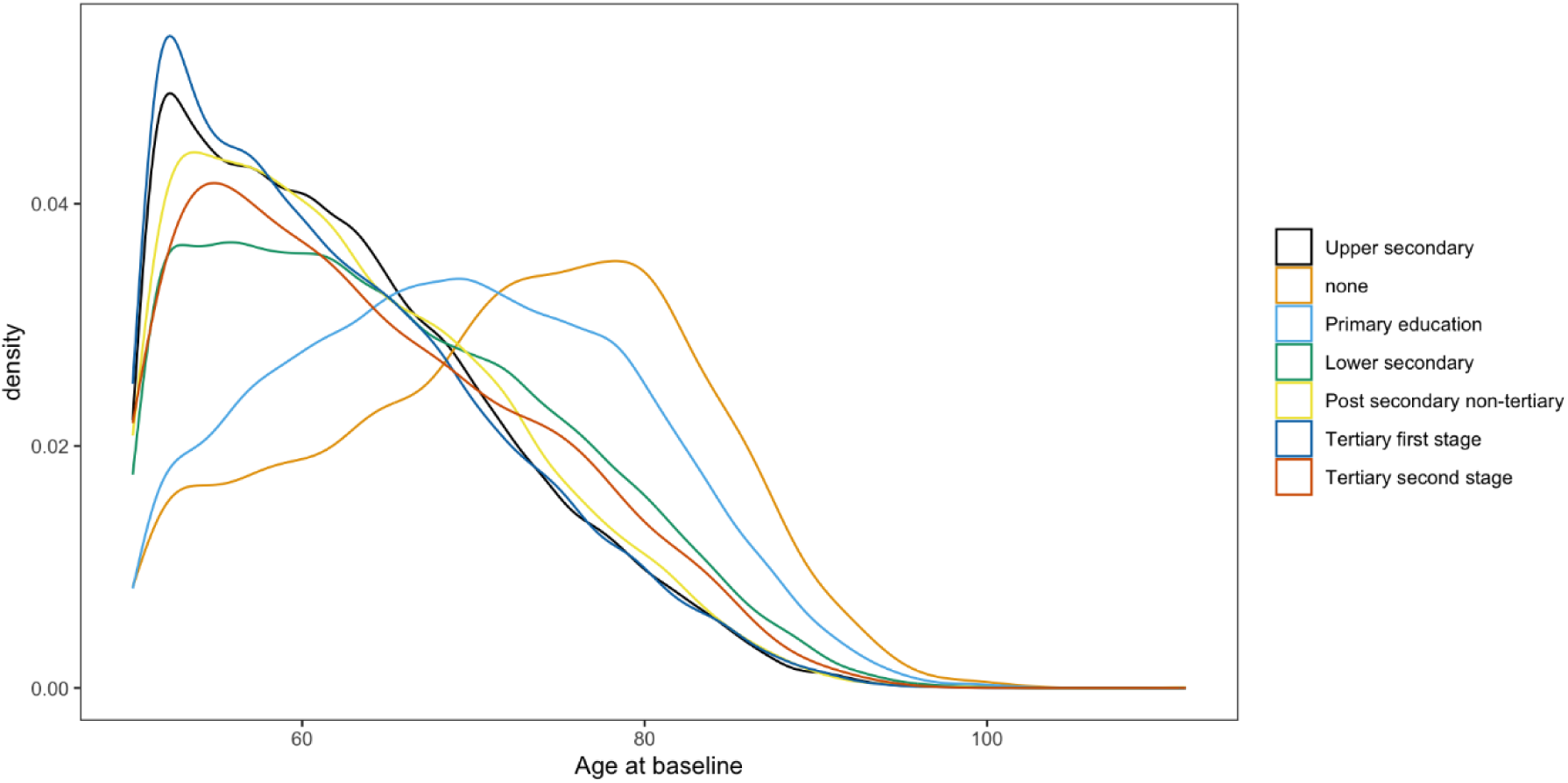
**Sample descriptives**. A: Number of participants for each test wave (not cumulative). B. Number of participants per education category. C. Age-distribution at baseline for each education category.

Memory was assessed with a 10 word verbal recall test. The word list is read out load to the participants, and then recall is tested immediately after the presentation (Recall 1) and then after a delay of approximately 5 minutes (Recall 2). Multiple versions of the lists are assigned to the respondents ^39^. The response distribution is shown in Figure SI 2. As can be seen, there are no ceiling effects, which is important when assessing longitudinal change for the best-performing participants. There are some floor effects for recall 2, but less for recall 1, suggesting that we can estimate longitudinal chance well for most baseline levels of memory. Since education is association with differences in memory scores, ceiling and floor effects could potentially obscure real differences in change, but this is unlikely to have affected the current results given the response distribution below. Scores were lower for delayed than immediate recall (OR = 0.535, CI: 0.534 – 0.537) and females scored higher than males (OR = 1.160, CI: 1.153-1.168).

**Online Figure:**
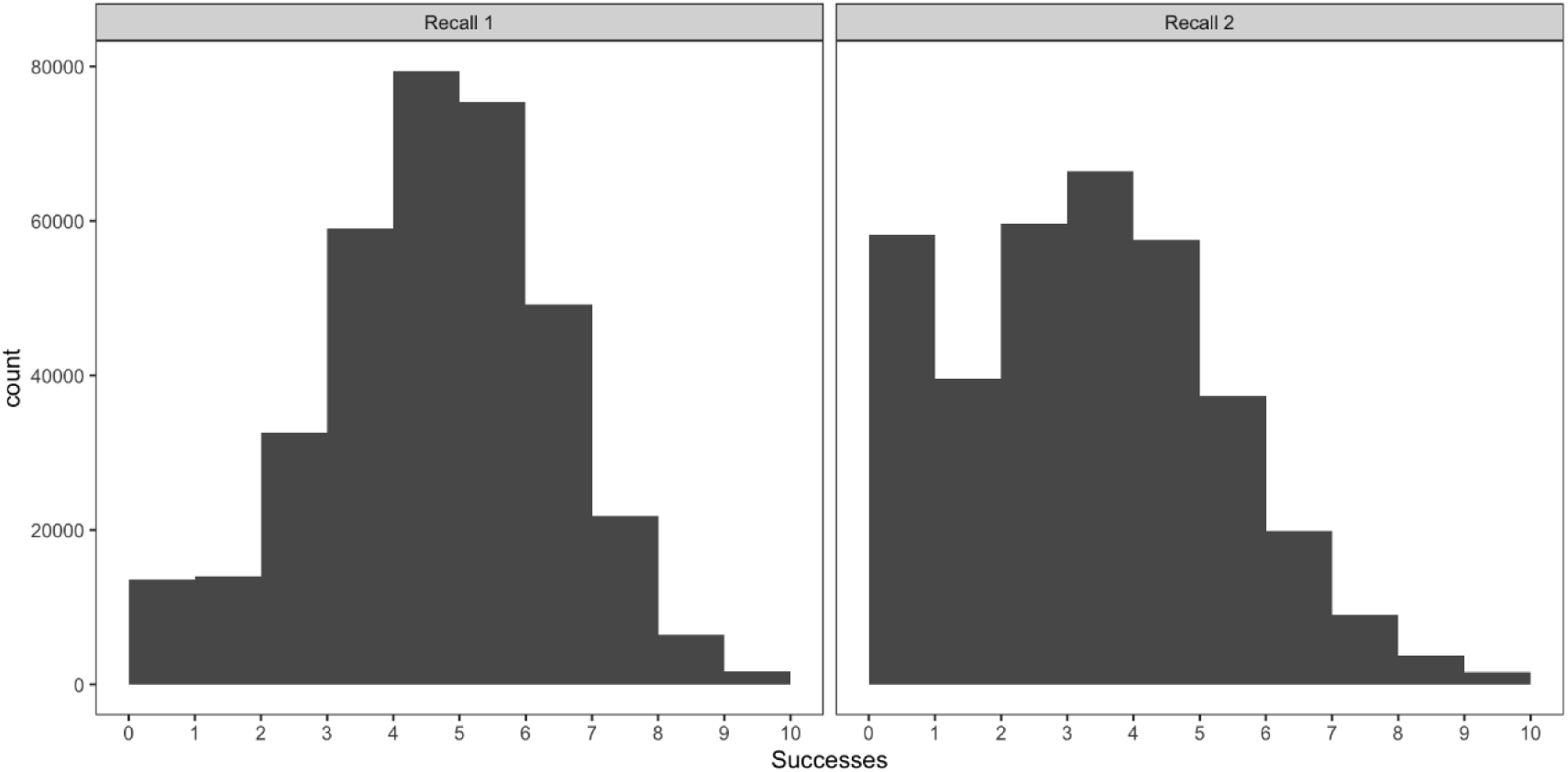
**Response distribution for word recall**. Number of participants (y-axis) as a function of number of words recalled (x-axis). Left: Immediate recall (Recall 1). Right: Delayed recall (Recall 2).

In addition to the memory measures, we extracted the variables age, sex, birth year, education (based on the International Standard Classification of Education 1997), and country of current residency.

### Statistical analyses: SHARE

Analyses were performed in R version 4.4.1 ^73^ using the brms package’s ^74^ interface to the probabilistic programming language Stan ^75^. To assess effects of education on memory and memory change, we ran logistic regressions with memory recall as dependent variable, yielding odds ratios as the most relevant model parameter to interpret. An odds ratio of 1 corresponds to a regression coefficient of 0. The main model was:

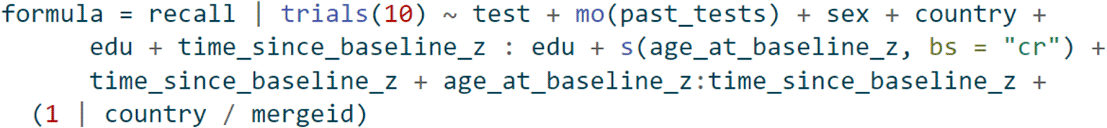

Each memory test was used as a separate response, yielding two observations per timepoint, and the variable *test* represents difficulty of condition 2 relative to condition 1. To control for practice effects, a monotonic function of the number of previous tests taken was included as covariate. We used a smooth function of age to allow non-linear relationships. Individual-specific intercepts per participant were nested within country. Default priors were used for all parameters, two parallel chains of Stan’s No-U-Turn Sampler ^76^ were run for 1500 iterations, discarding the first 1000 as warmup. This yielded 1000 post-warmup samples. For the offset/level analyses, education (edu) was the variable of interest, while for the slope/change analyses, edu × time since baseline was the critical variable. Z-transformed variables were used in the model fitting for numerical stability, and results converted back to their natural units for easier interpretability, e.g., age and time in years.

#### Brain cohorts

We combined data from 13 datasets with longitudinal brain MRIs and memory assessments: LCBC ^77^, Betula ^78,79^, UB ^80,81^, BASE-II ^82,83^, and Cam-CAN ^84^ datasets (from the Lifebrain Consortium) ^38^ as well as the COGNORM ^85^, the Alzheimer’s Disease Neuroimaging Initiative (ADNI) database (https://adni.loni.usc.edu) ^86^, BBHI ^87^, the Harvard Aging Brain Study (HABS) ^88^, the UKB (https://www.ukbiobank.ac.uk/) ^89^, PREVENT-AD ^90,91^, OASIS3 (https://sites.wustl.edu/oasisbrains/) ^92^, and VETSA ^93^. Sample size was maximized for each analysis and hence varies due to data availability and missingness (see Table 1 for an overview). In addition to cohort-specific inclusion and exclusion criteria, participants >50 years without cognitive impairment, Alzheimer’s dementia or severe neurological or psychiatric disorders were included. Additionally, MRI data from scanners with fewer than 15 measurements were also excluded. The initial dataset included individuals with 1 to 14 MRI acquisitions with longitudinal structural MRI data spanning up to 15.8 years. Similarly, memory assessments range from 1 to 24 observations per individual with a follow-up up to 28 years. For detailed descriptions of general characteristics of each dataset, please refer to the study-specific citations above. A general overview of each dataset is given in the table below.

**Online Table:**
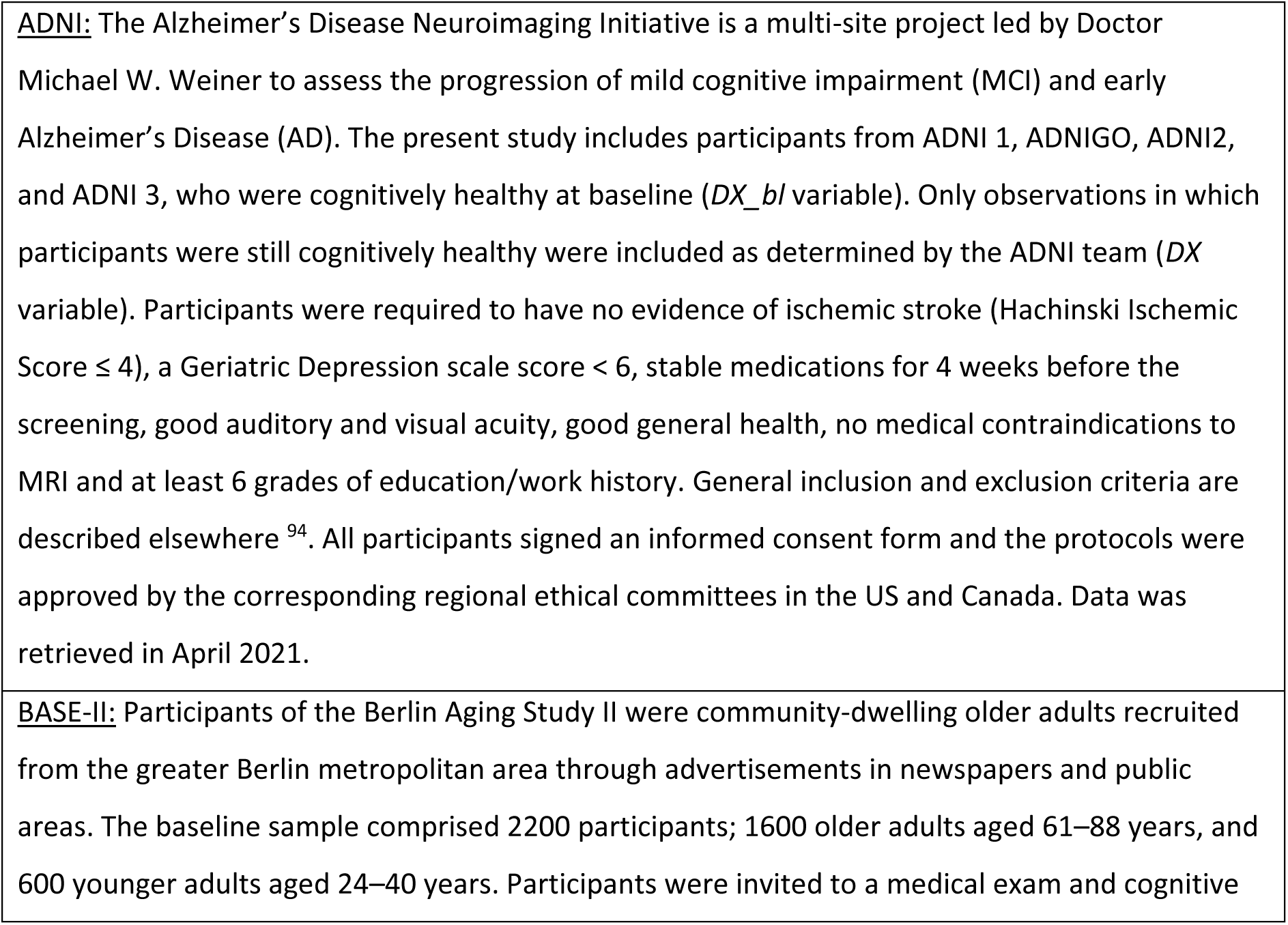
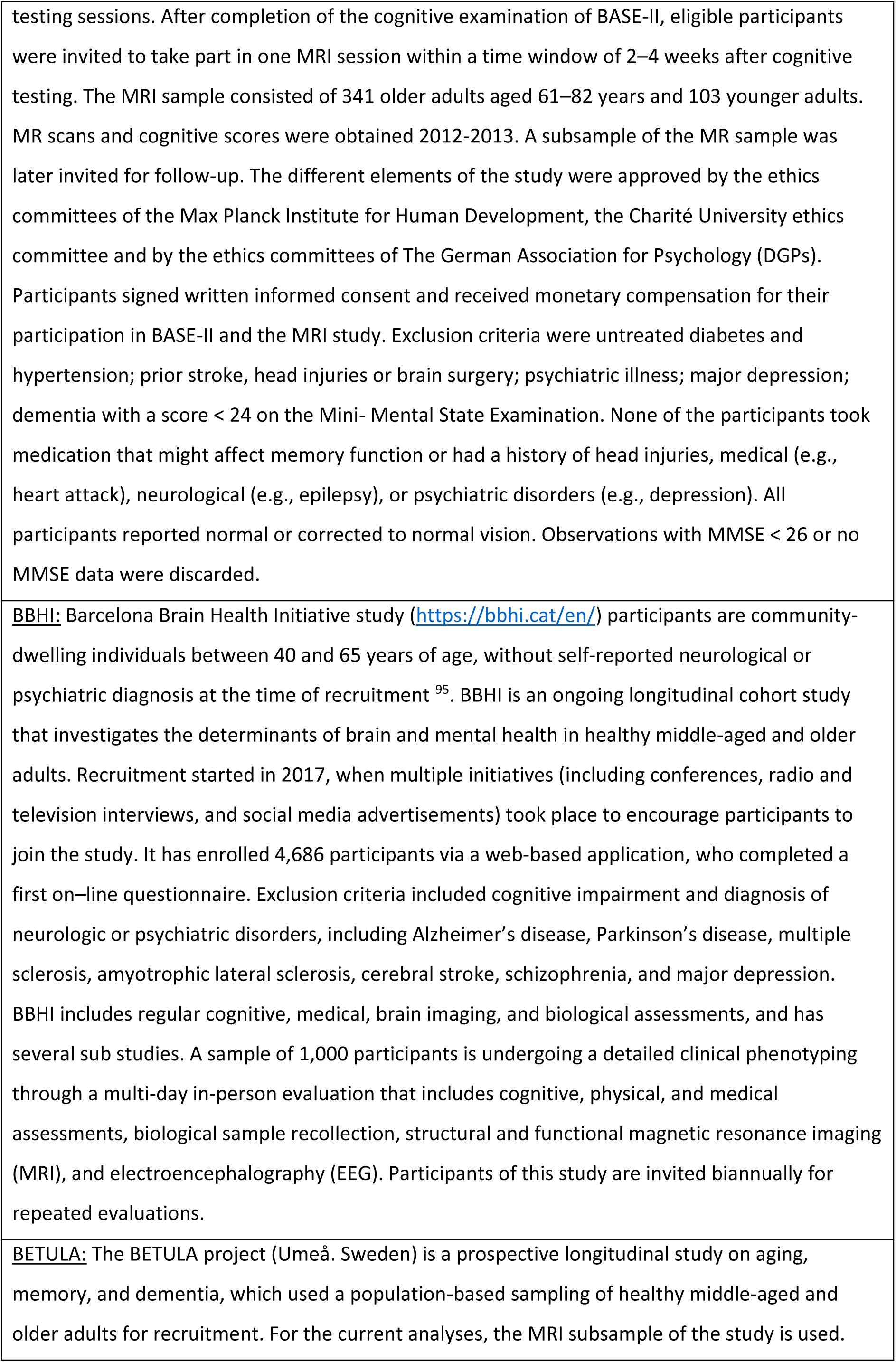
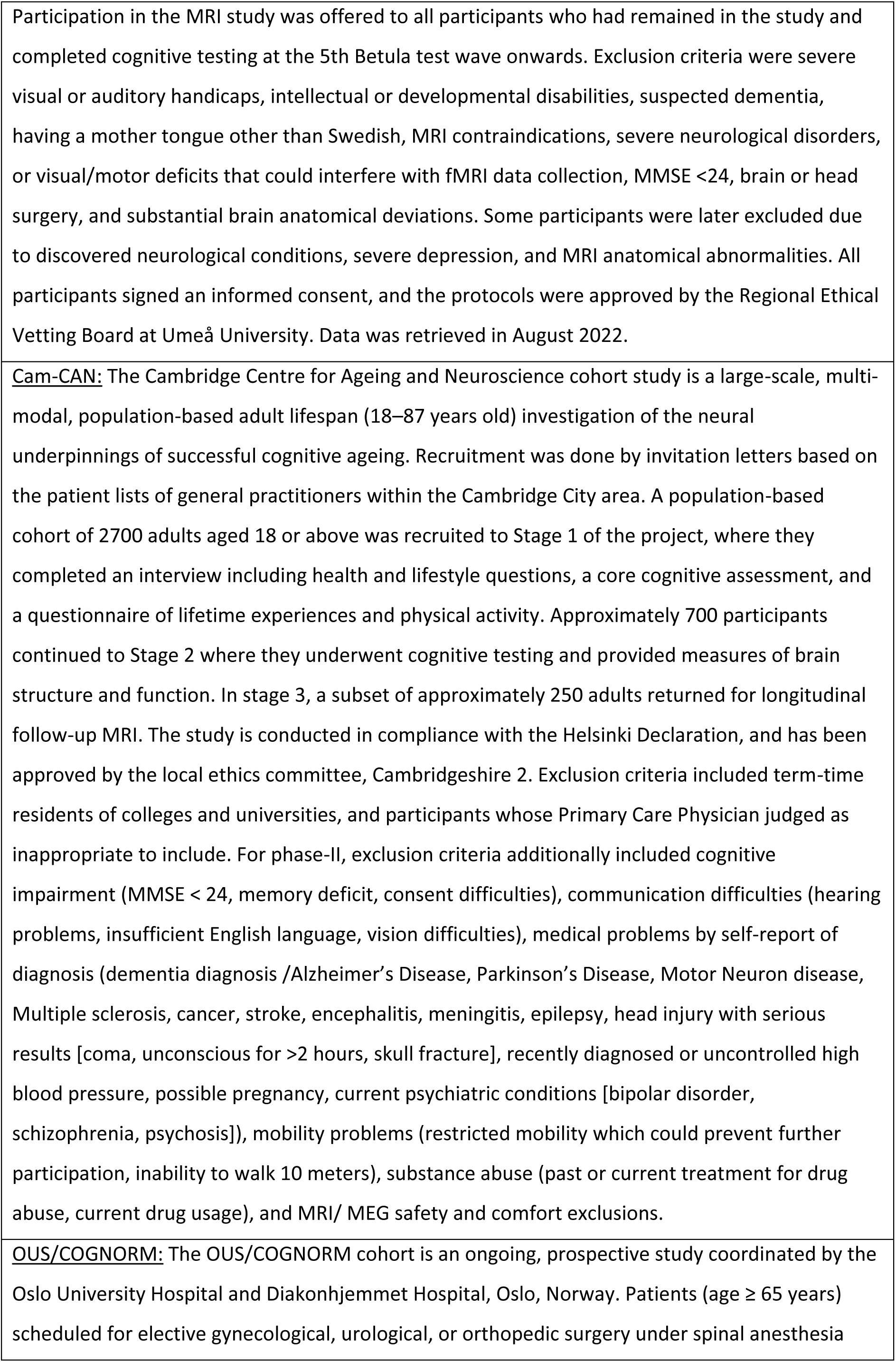
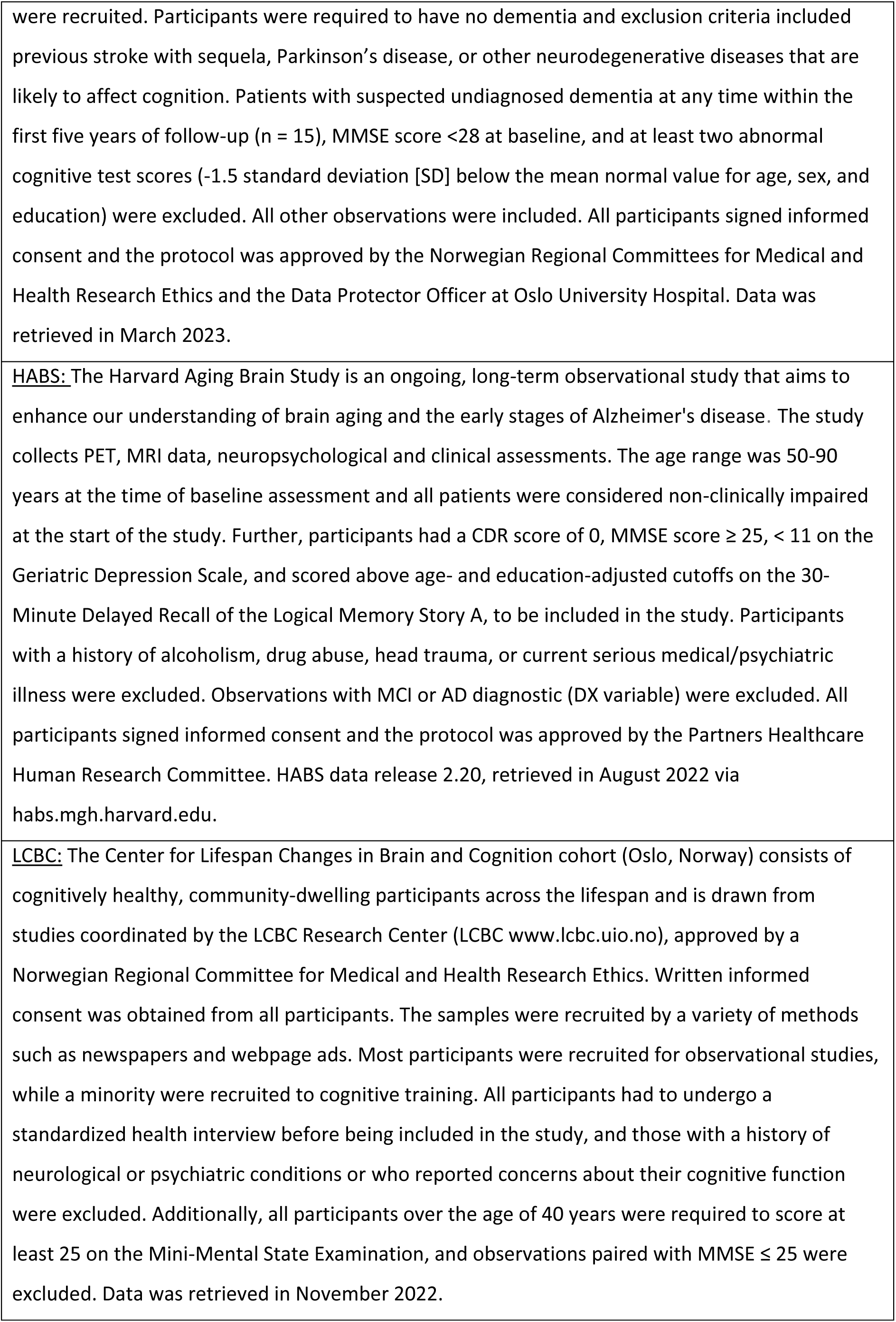
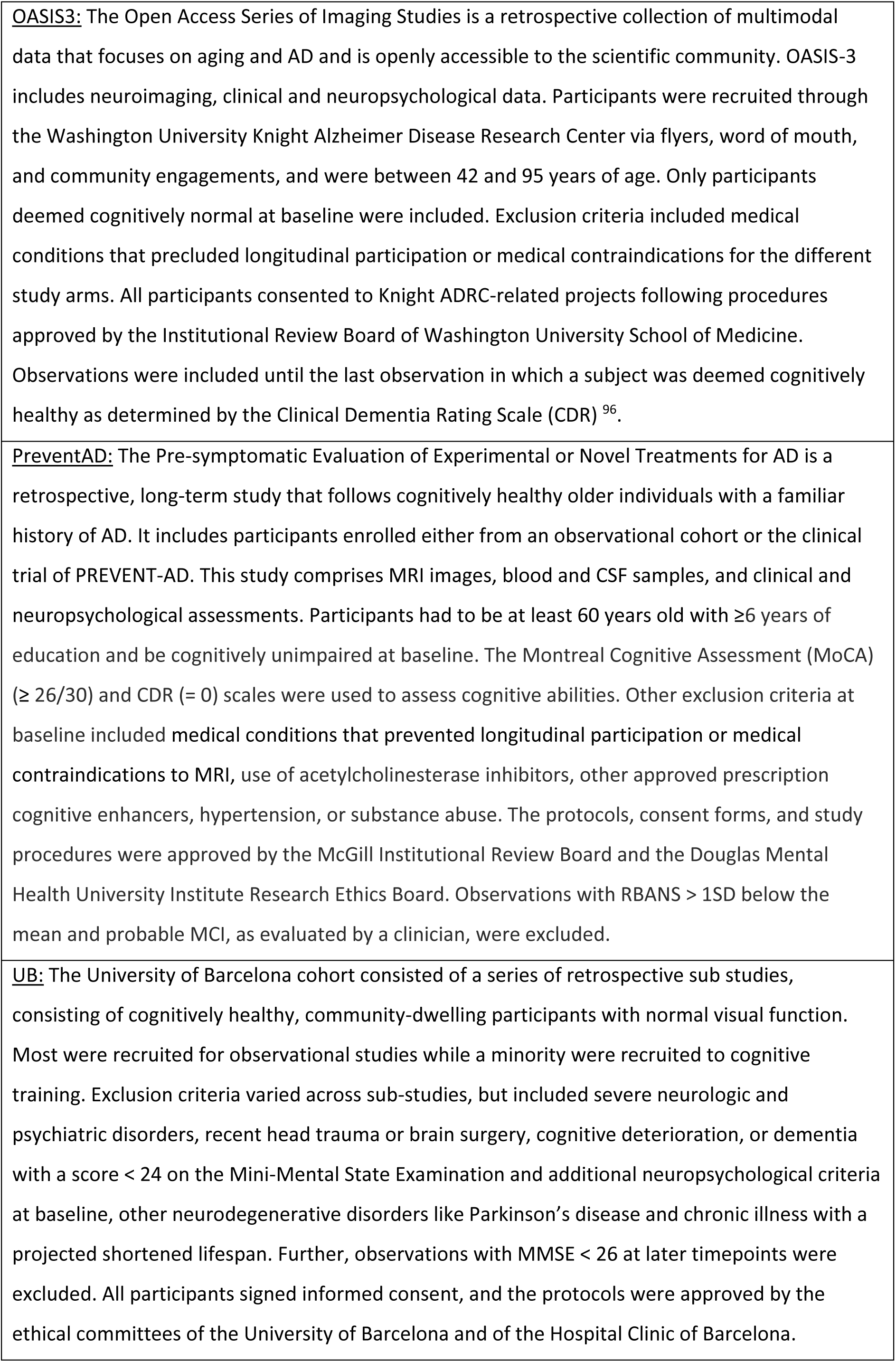
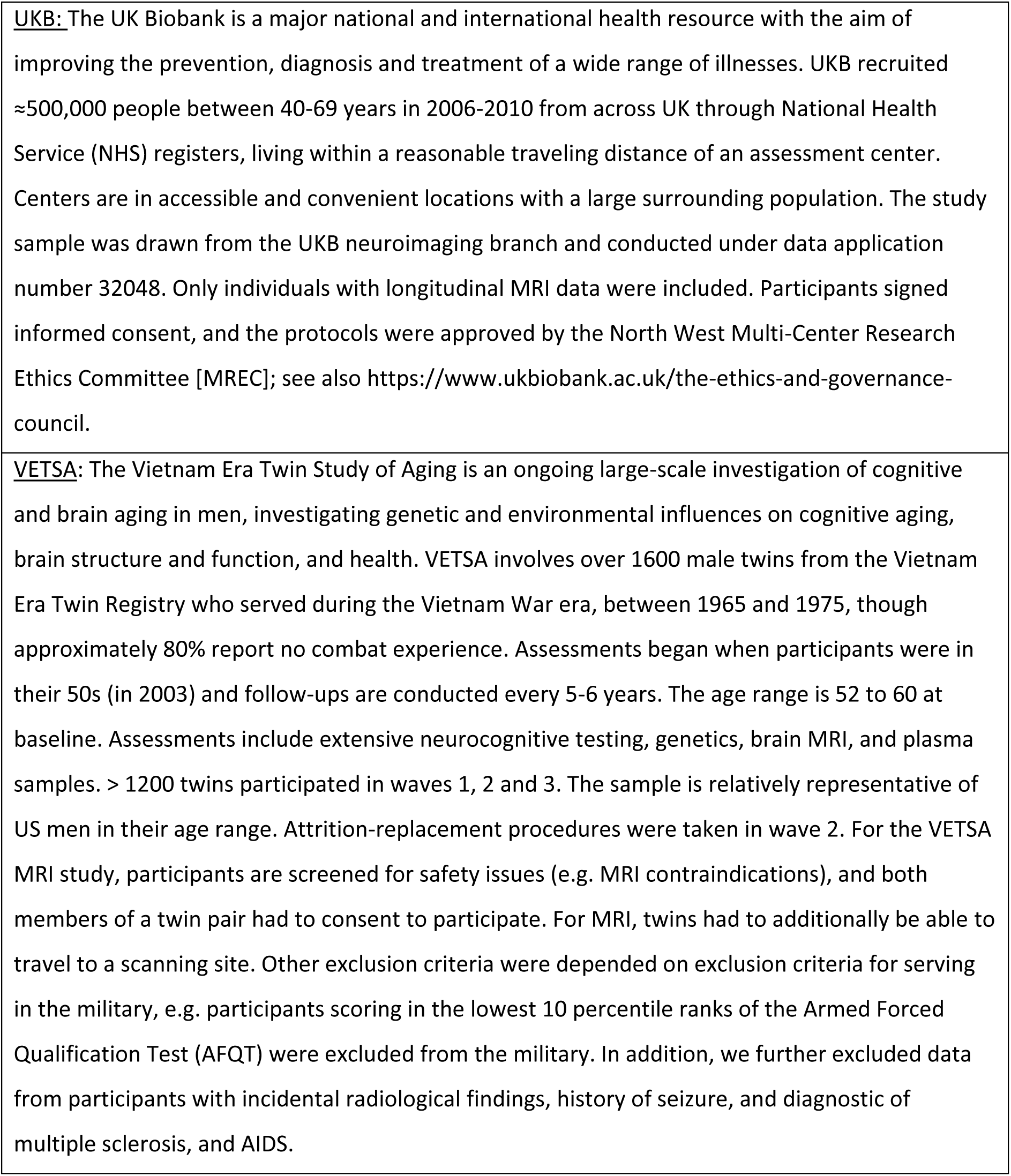
General characteristics of each sample.

The main sample descriptives are provided in Table 1 in the main manuscript. Since the exact sample size varies between analyses depending on data availability, the specific characteristics for the samples used to address the different research questions are provided in the table below and the sample distributions shown in the figure.

**Online Table:**
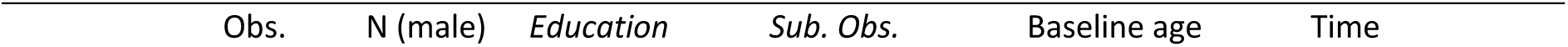
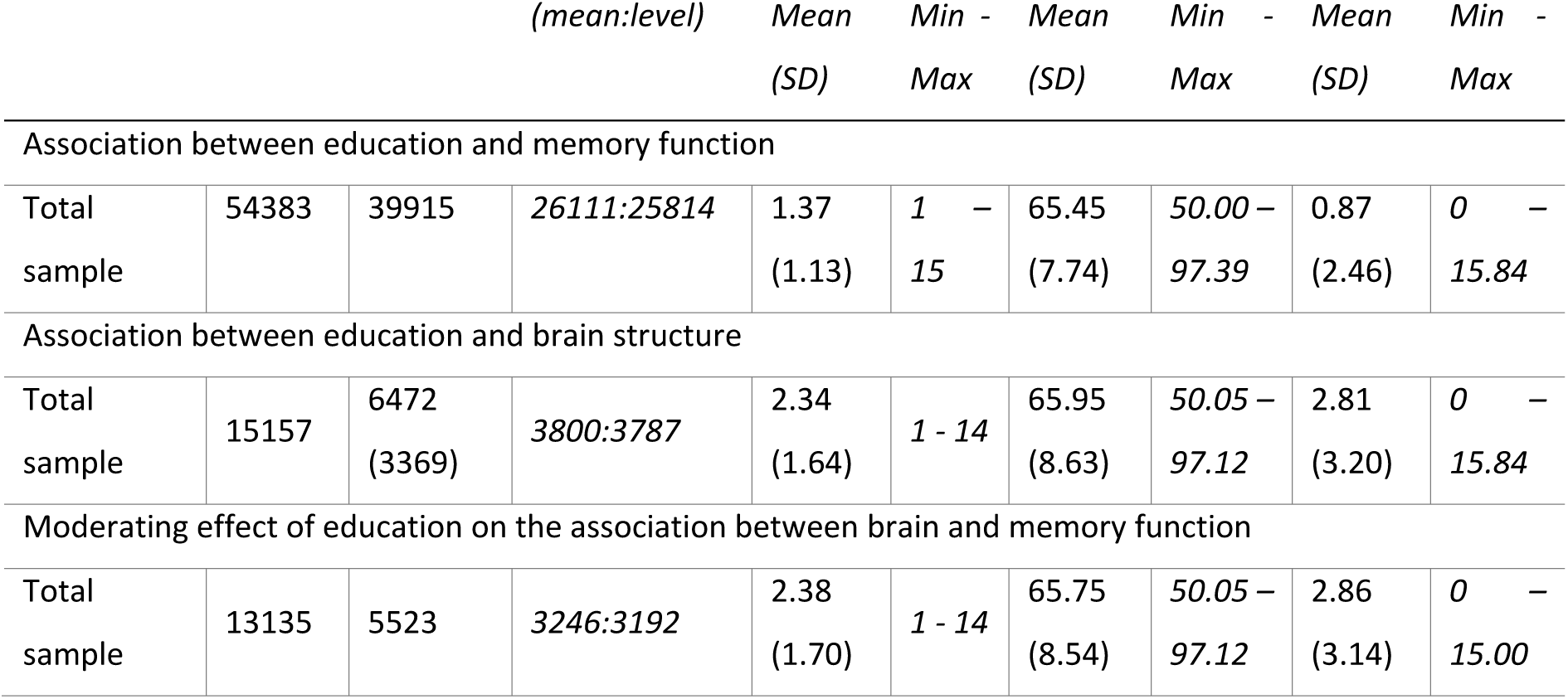
Main characteristics for the total dataset used to address the different classes of research questions.

| Obs. | N (male) | Education | Sub. Obs. | Baseline age | Time |
| --- | --- | --- | --- | --- | --- |

|  |  |  | (mean:level) | Mean<br>(SD) | Min -<br>Max | Mean<br>(SD) | Min -<br>Max | Mean<br>(SD) | Min -<br>Max |
| --- | --- | --- | --- | --- | --- | --- | --- | --- | --- |
| Association between education and memory function |  |  |  |  |  |  |  |  |  |
| Total sample | 54383 | 39915 | 26111:25814 | 1.37<br>(1.13) | 1 - 15 | 65.45<br>(7.74) | 50.00 - 97.39 | 0.87<br>(2.46) | 0 - 15.84 |
| Association between education and brain structure |  |  |  |  |  |  |  |  |  |
| Total sample | 15157 | 6472<br>(3369) | 3800:3787 | 2.34<br>(1.64) | 1 - 14 | 65.95<br>(8.63) | 50.05 - 97.12 | 2.81<br>(3.20) | 0 - 15.84 |
| Moderating effect of education on the association between brain and memory function |  |  |  |  |  |  |  |  |  |
| Total sample | 13135 | 5523 | 3246:3192 | 2.38<br>(1.70) | 1 - 14 | 65.75<br>(8.54) | 50.05 - 97.12 | 2.86<br>(3.14) | 0 - 15.00 |

**Online figure:**
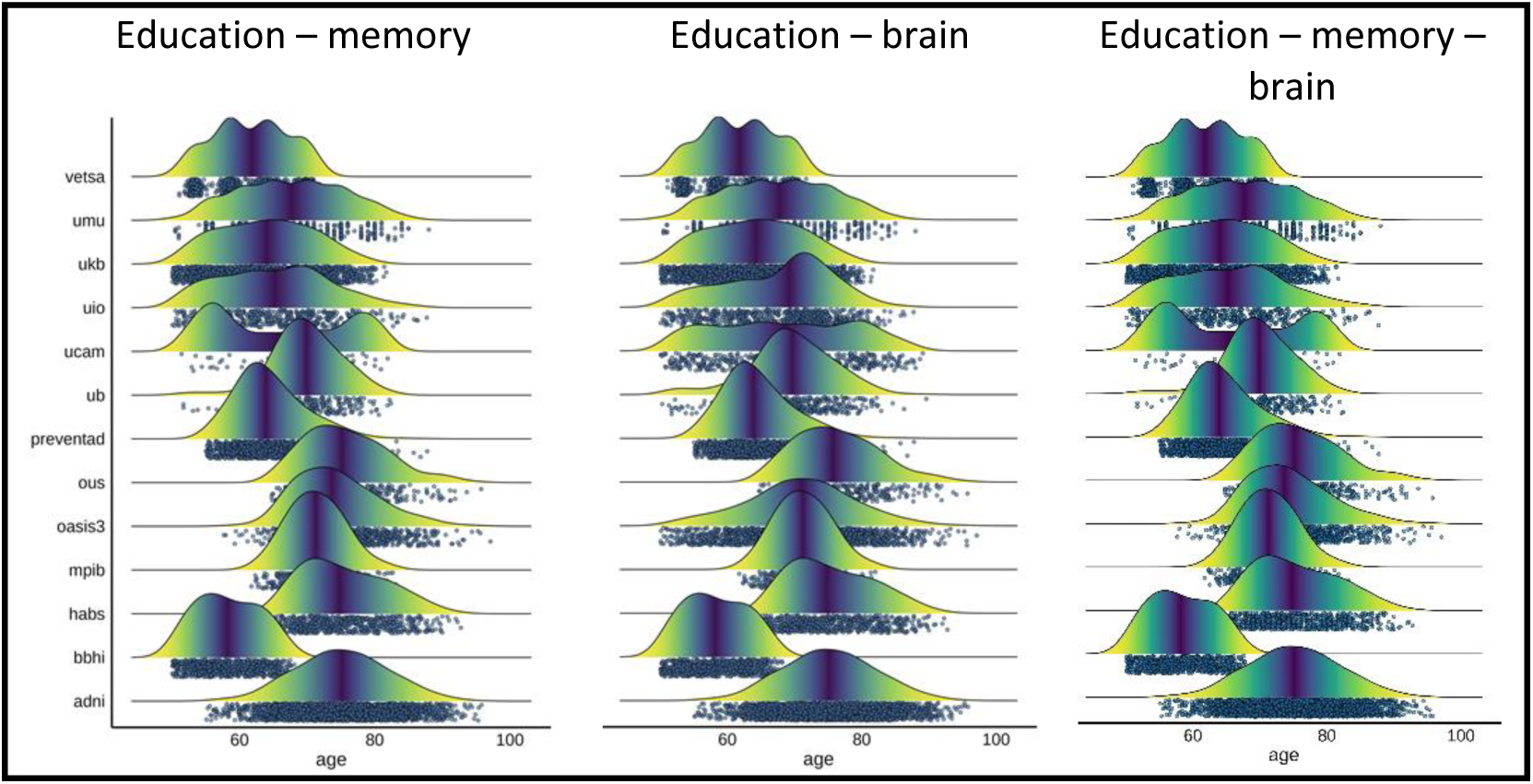
**Sample distribution of the brain cohorts used for testing education-memory (left), education-brain (middle) and education-memory-brain relationships (right).**

### Data availability

Each dataset has different owners. Contact information to be used for data is specified in the table below.

**Online Table:**
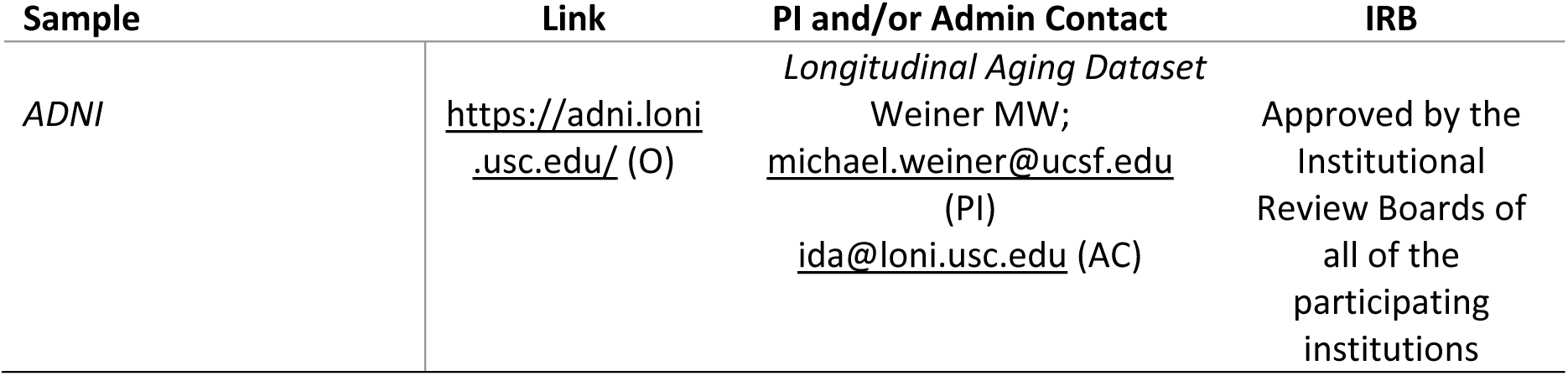
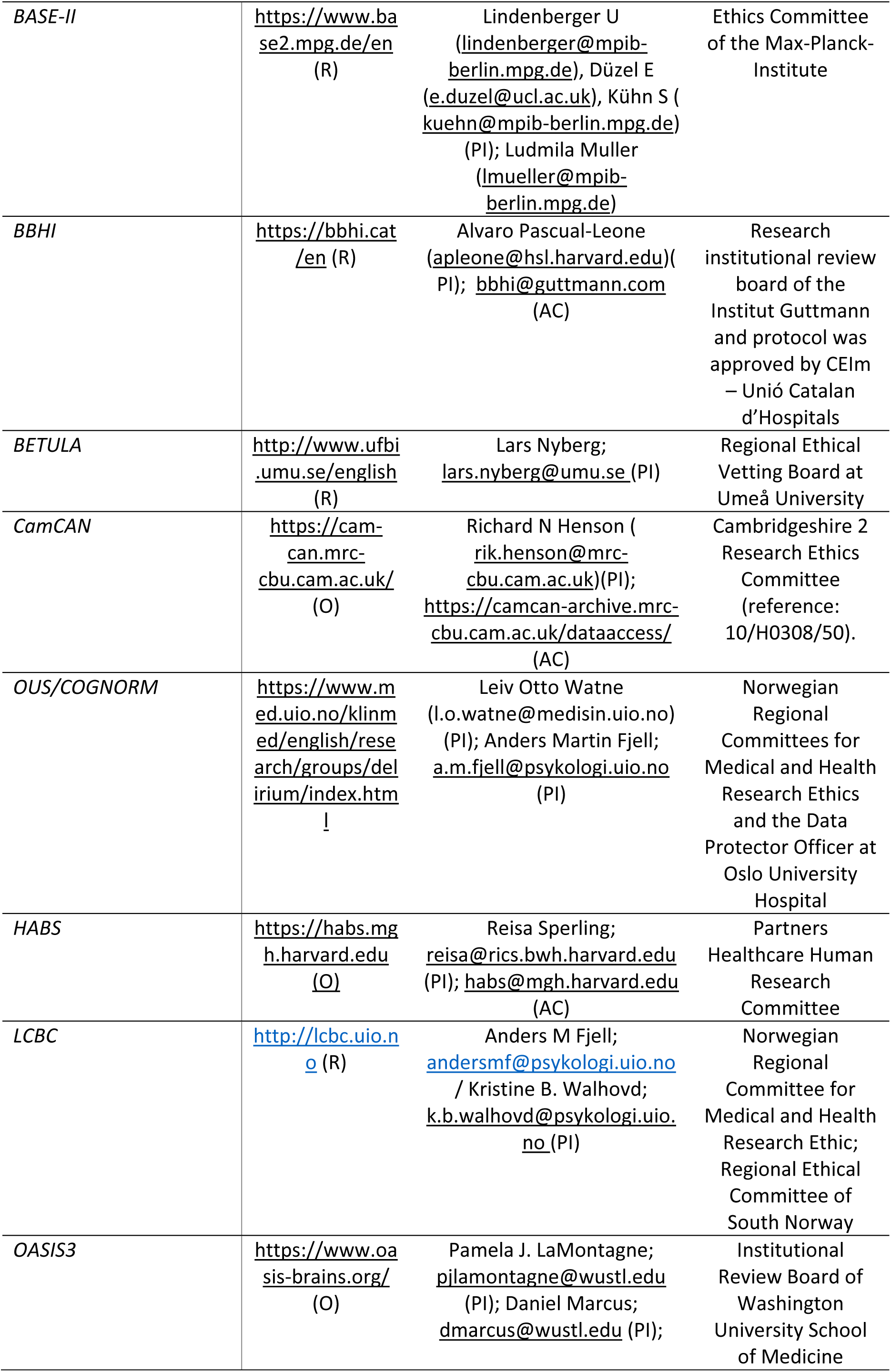
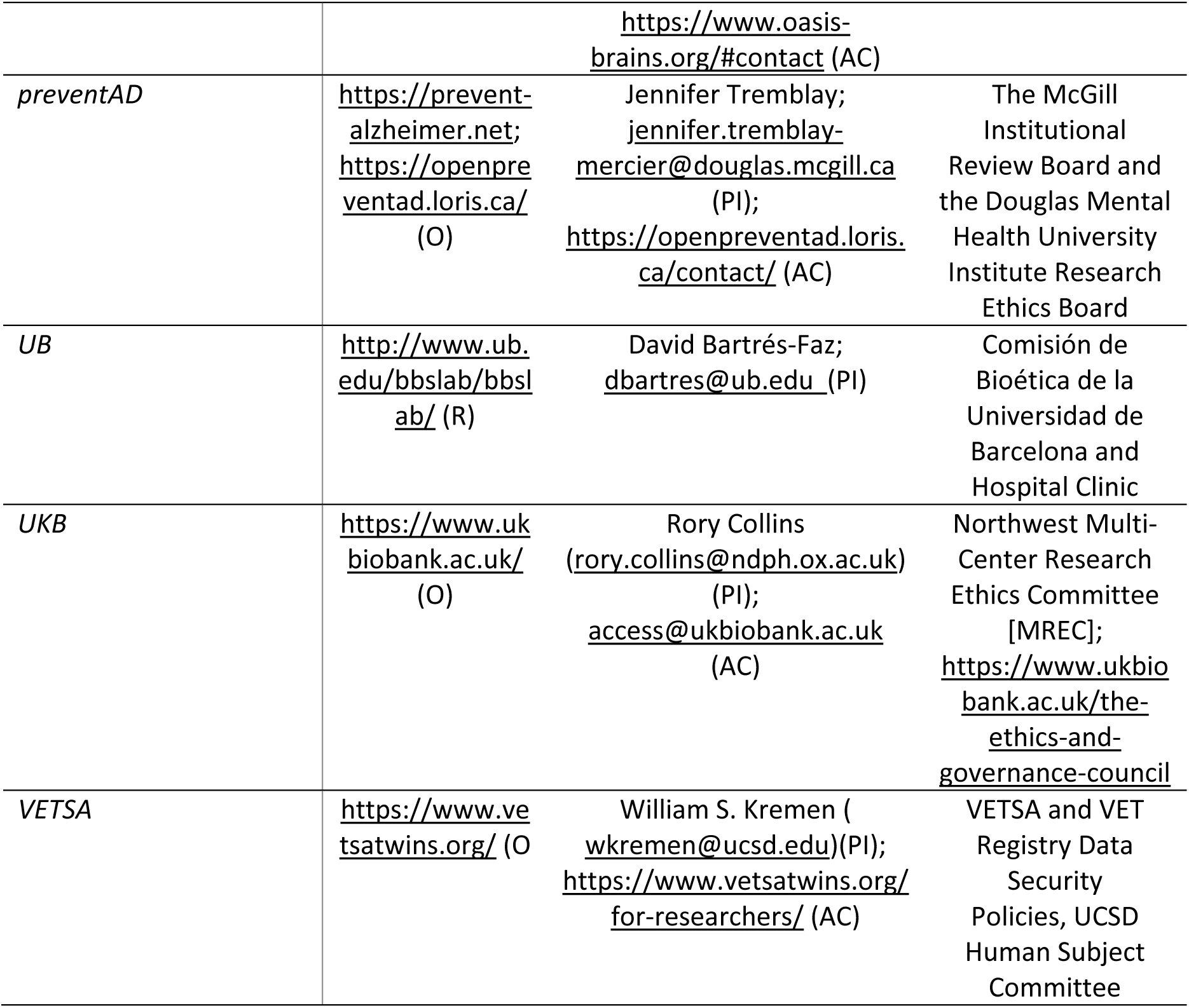
Links, data owner and IRB approvals for each dataset. Data availability, contact and principal investigator information, and ethical approval for the different datasets used. PI = Principal Investigator. AC = Administrative contact. IRB = Institutional Review Boards. O = Openly available. Automatic or semi-automatic data agreements. Fees may apply (e.g. UKB). R = Restricted. Ad-hoc permission is required. Contact PI or AC for specific details on access to data.

### Education in the brain imaging cohorts

For each dataset, education was categorized as high or low using a mean split. We chose this approach because quantitative distributions of education were often highly non-gaussian and level-based codifications were somewhat arbitrary due to idiosyncratic reporting of years of education, and variations in schooling systems across years and country. To ensure robustness, we conducted analyses with an alternative operationalization of education, categorizing individuals with or without tertiary education. When education data was provided as qualifications or categories, these were converted to years of education based on country-specific norms. Individuals were then grouped as having high or low education based on the median. For the tertiary education categorization, the reverse process was applied, converting years of education into education qualifications. For reporting consistency, a lower cap of 6 years and an upper cap of 20 were applied to education years. An overview of education characteristics is provided in the table below and visualized in the figure.

**Online table:**
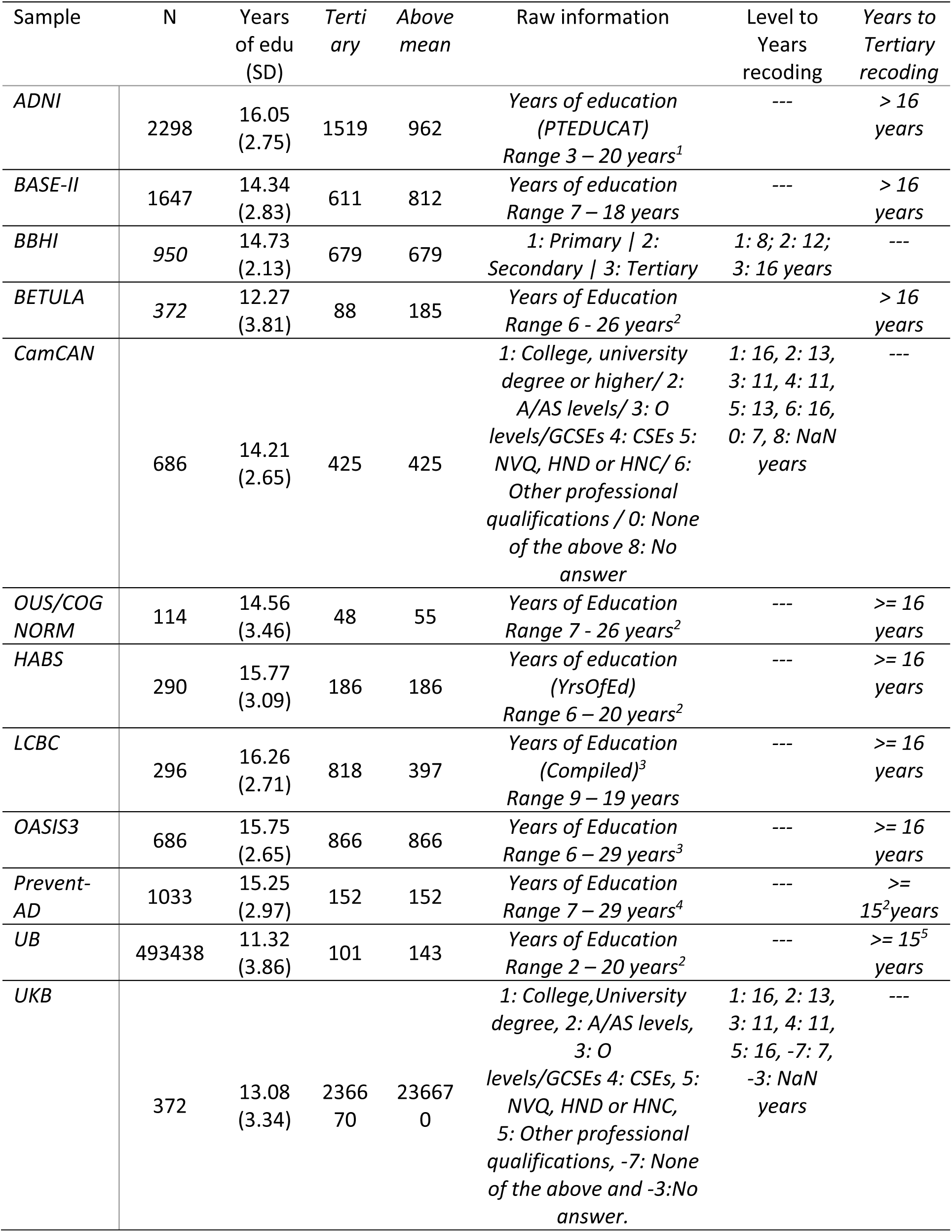
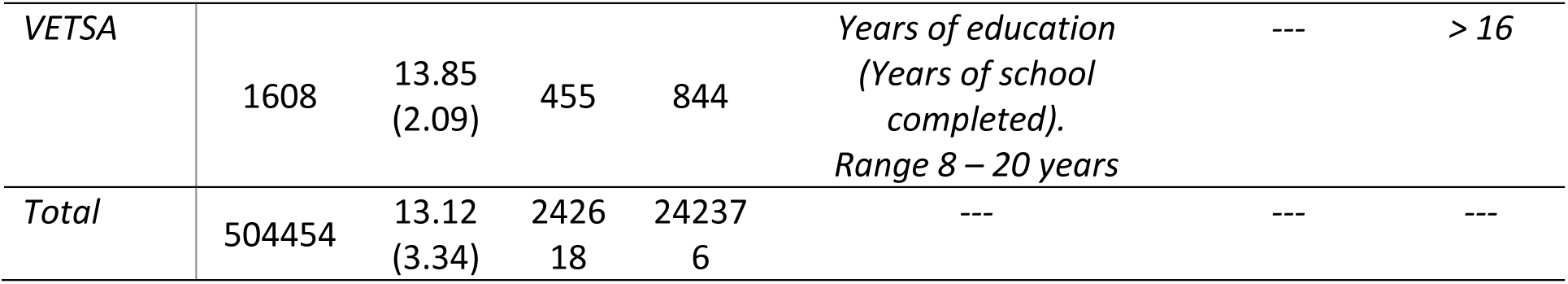
Overview of education variables and recoding. Education data from the MRI sample. ^1^Lower cap at 6 years. ^2^Capped at 20 years. ^3^Different sources. Converted to semi-quantitative values with 9, 12, 16, and 19 years of education corresponding to basic, secondary, tertiary, and upper tertiary education. ^4^Based on Quebec norms (https://www.quebec.ca/en/education/study-quebec/education-system). ^5^Education system changes throughout through the 20th century in Spain varies the minimum years of education required to acquire tertiary education.

**Online figure:**
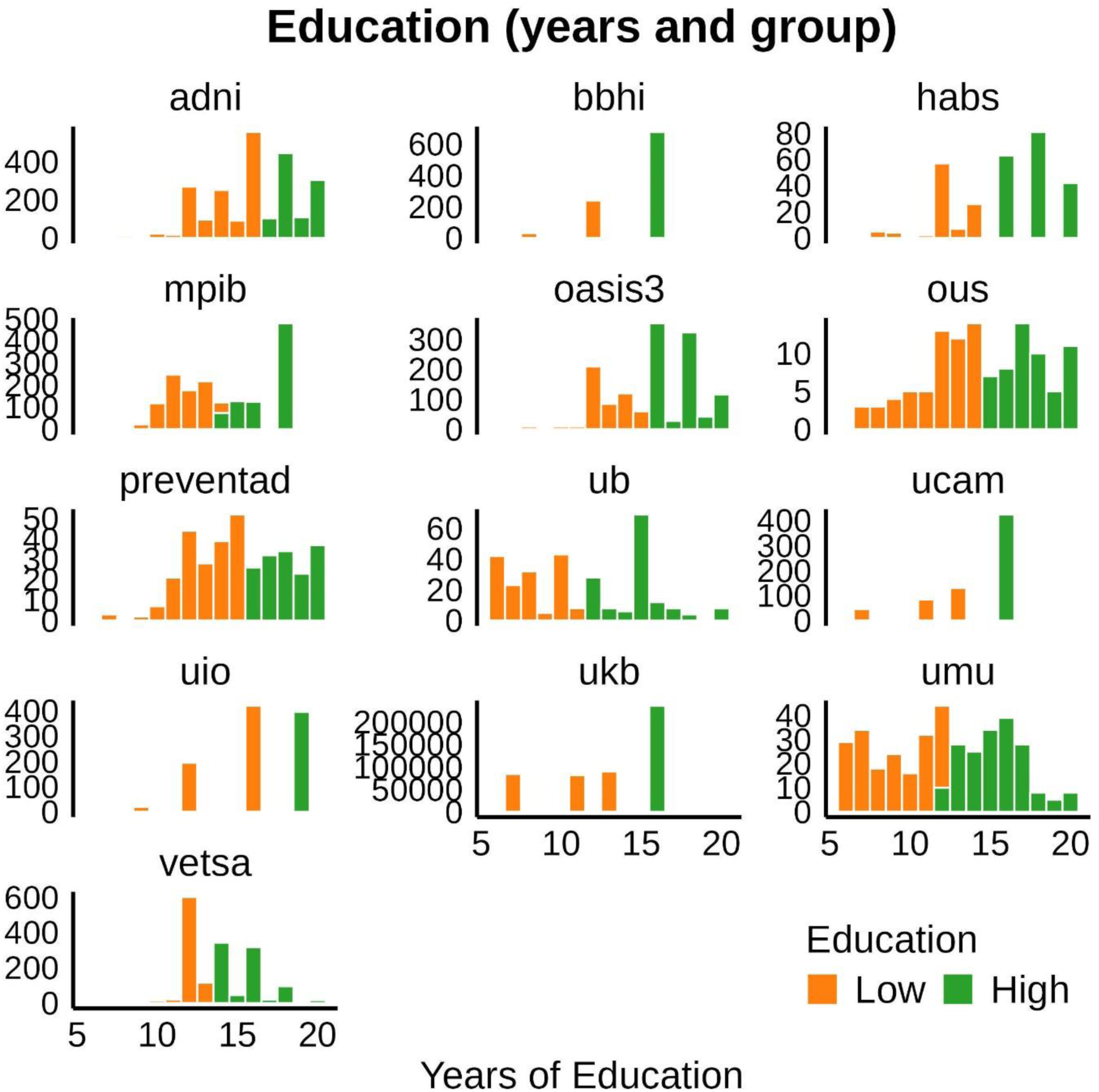
**Distribution of education in each sample**

### Memory function in the brain imaging cohorts

For each sample, we operationalized memory performance as a z-normalized score based on the first time point and the different available memory tests. When multiple scores were available, we used the first component of a Principal Component Analysis (PCA) with all measures as inputs. For each dataset, we regressed out age (as a smoothing term), sex, and one or two dummy test-retest regressors using generalized additive mixed models (*gamm4 R-package*) ^40^. Individual identifiers were used as random intercepts and the number of dummy test-retest regressors depended on whether the dataset had 2 or >=3 waves with memory function data. The residuals were used as an estimate of memory function in each observation. An overview of tests included in the memory performance score for each dataset is provided in the table.

**Online table:** Tests related to episodic memory included in the analyses for each sample. A PC was estimated based on the first time point for which multiple memory measures were available. Participants (Obs.). Participants and Observations with memory from the initial mri sample. MMSE = Mini-mental State Examination. RAVLT = Rey Auditory Verbal Learning Test; CVLT = California Verbal Learning Test; PAL = Paired associate learning (#20197 UKB field); Logical memory = Memory subtest of the Wechsler Memory Scale. CERAD = Consortium to Establish a Registry for Alzheimer’s Disease (CERAD) Word List Memory test. ADAS = Alzheimer Disease Assessment Scale. VMLT = Verbal Learning and Memory test. SRT = Buschke Selective Reminding Task. RBANS = Repeatable Battery for Assessment of Neuropsychological Status. Story recall = Story recall and recognition task of episodic memory from Wechsler Neuropsychological Battery. ^1^ADNI-MEM score was computed developed by ^97^ and consists of a composite score of memory which includes measures from RAVLT (learning trials, list, recognition and recalls), ADAS (learning trials, recall, and recognitions), MMSE words, and Logical memory. ^2^See Nilsson et al^98^ ^3^Second wave was administered online. Calibration data (not shown here) shows in person vs. online data is comparable. ^4^See https://biobank.ndph.ox.ac.uk/showcase/refer.cgi?id=2561 for more information on PAL.

| Dataset | Participants (Obs.) | Memory Tests |
| --- | --- | --- |
| ADNI | 904 (3824) | ADNI-MEM<sup>1</sup> |
| BASE-II | 1894 (3110) | VLMT short delay recall<br>VLMT long delay recall<br>VLMT learning (sum across trials) |
| BBHI | 966 (1266) | RAVLT learning (sum across trials)<br>RAVLT long delay recall<br>RAVLT short delay recall |
| BETULA | 337 (1563) | Recall of sentences<sup>2</sup> |
| CamCAN | 89 (172) | Story short delay recall<sup>3</sup><br>Story long delay recall |
| OUS/COGNORM | 114 (667) | CERAD short delay recall<br>CERAD long delay recall |
| HABS | 287 (1289) | Logical memory short delay recall<br>Logical memory long delay recall<br>SRT delayed recall<br>SRT total recall |
| LCBC | 938 (1440) | CVLT short delay recall<br>CVLT long delay recall<br>CVLT learning (sum across trials) |
| OASIS3 | 648 (3170) | Logical memory immediate |
| preventAD | 306 (1057) | RBANS list recall<br>RBANS list learning (sum across trials)<br>RBANS story immediate memory<br>RBANS story delayed recall |
| UB | 161 (298) | RAVLT learning (sum across trials)<br>RAVLT long delay recall |
| UKB | 33,890 (36,520) | PAL<sup>4</sup> |
| VETSA | 1592 (3617) | CVLT short delay recall<br>CVLT long delay recall<br>CVLT learning (sum across trials) |

### Magnetic Resonance Imaging acquisition and preprocessing

Structural T1-weighted (T1w) MPRAGE and FSPGR scans were collected using 1.5 and 3T MRI scanners. Information regarding scanners and scanner parameters across datasets are presented in the table below.

**Online table:**
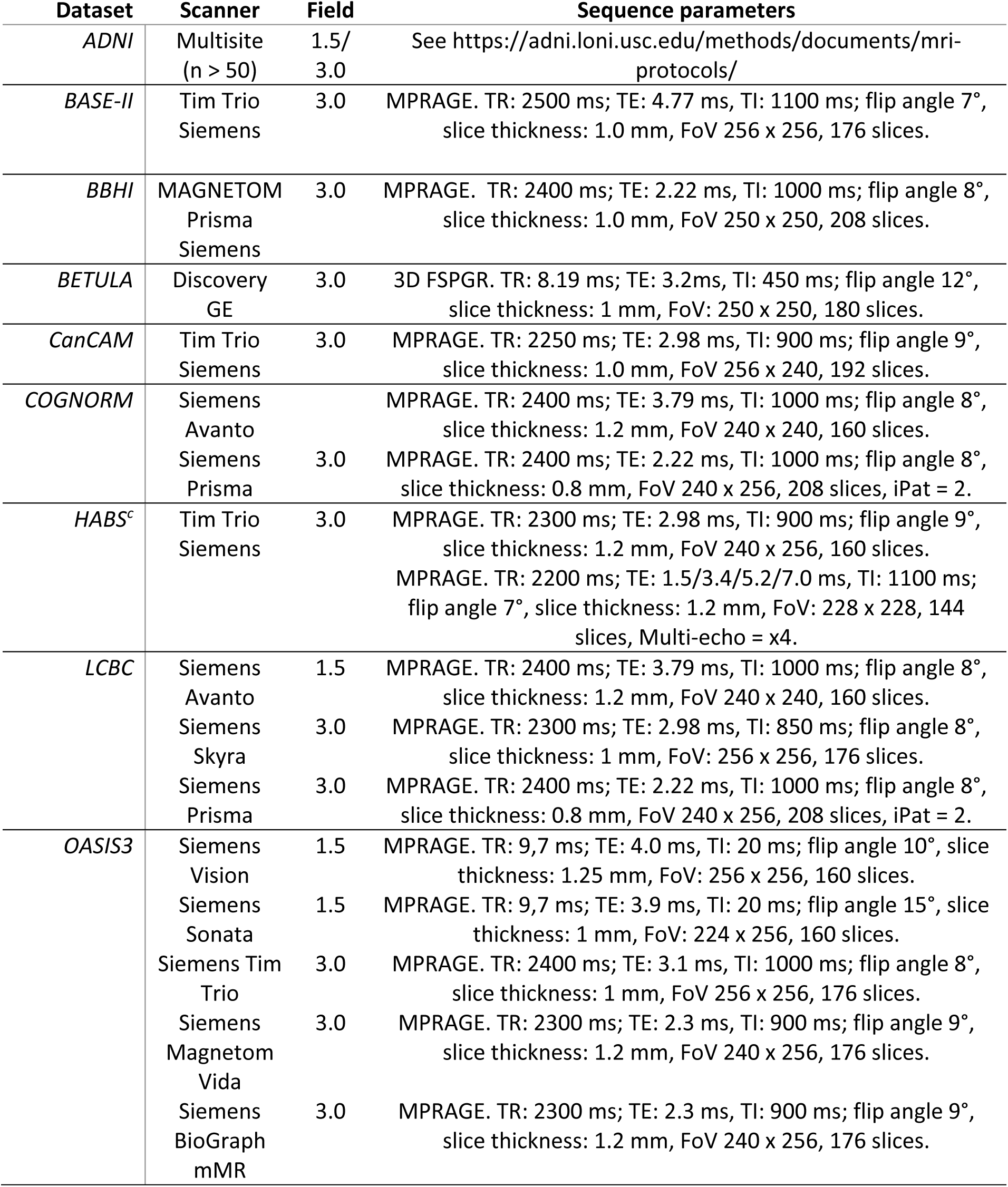
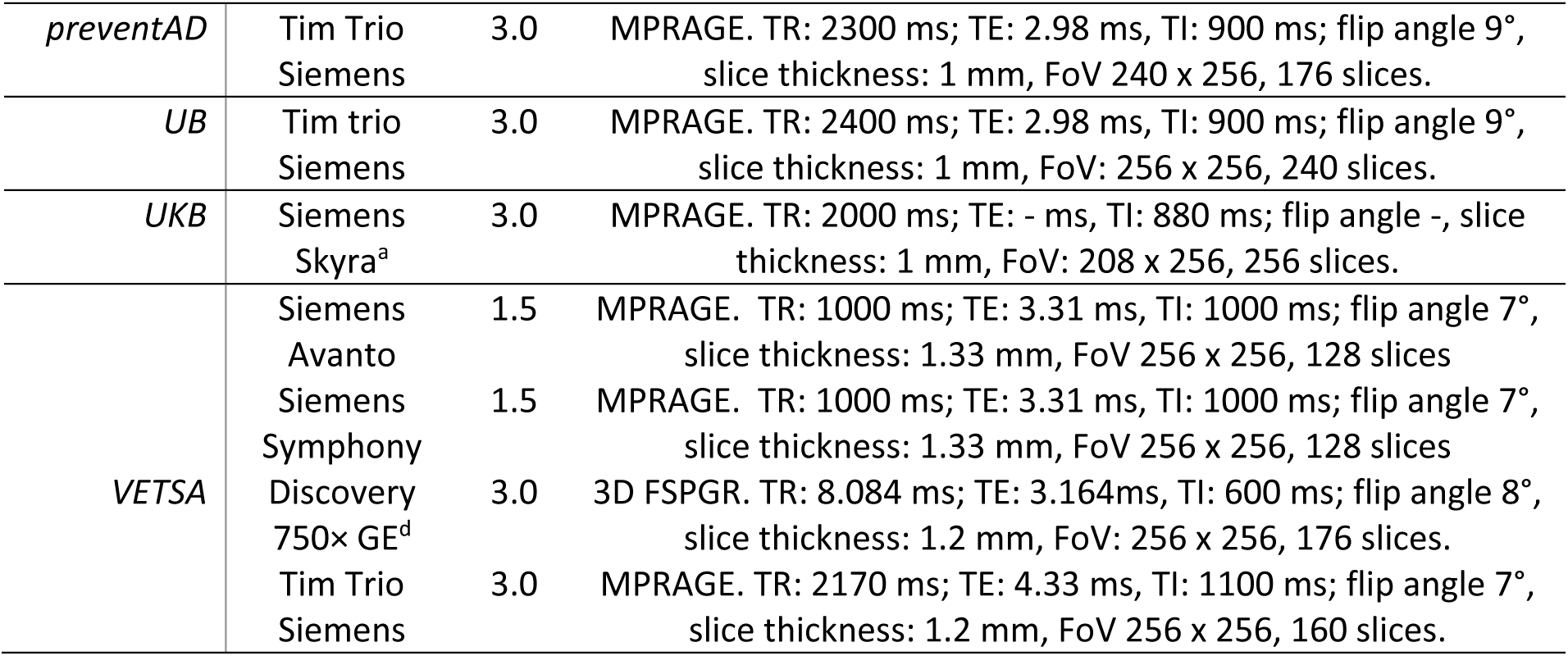
Scanner acquisition parameters. TR = Repetition Time; TE = Echo Time; TI = inversion time; FoV = Field of View, iPat = in-plane acceleration. ^a,c,d^Two matched scanners. ^b^Several matched scanners.

For datasets not provided in Brain Imaging Data Structure (BIDS) format, data was converted to BIDS^99^. BIDS transformation of ADNI, OASIS3, and HABS data were performed with Clinica software ^100,101^. We used the longitudinal FreeSurfer v.7.1.0 stream ^102^ for cortical reconstruction and volumetric segmentation of the structural T1w scans ^103–105^. For sessions with multiple scans, data from the scanners were averaged. Briefly, the images were processed using the cross-sectional stream, which includes the removal of nonbrain tissues, Talairach transformation, intensity correction, tissue and volumetric segmentation, cortical surface reconstruction, and cortical parcellation. Next, an unbiased within-subject template space based on all cross-sectional images was created for each participant, using robust, inverse-consistent registration (Reuter et al., 2010). The processing of each time point was then reinitialized with common information from the within-subject template, to increase reliability and statistical power. Except for the BETULA dataset, all data was preprocessed on the Colossus processing cluster, part of the Services for Sensitive Data (TSD) (https://www.uio.no/tjenester/it/forskning/sensitiv/), University of Oslo. Memory-sensitive brain measures for each observation were derived using regional loadings based on the *Destrieux* (cortical)^106^ and *aseg* (subcortical) atlases ^107^.

### Memory-sensitive brain measures

We computed two complimentary measures of brain structure sensitive to memory, capturing different aspects of memory function in older age. The primary measure was defined as a longitudinal brain component sensitive to memory changes inspired by Vidal-Pineiro et al. (*in preparation*). The second measure, for the purpose of assessing the robustness of the results, was trained on independent scans to detect cross-sectional brain-memory relationships in aging. The components were highly correlated (r = .71), both decrease with age (r = −.67, r = −.64, respectively) and include partially overlapping set of brain regions. The first measure (brain PC) is optimized to be sensitive to memory changes in aging, while the second (brain LASSO) is optimized to detect also offset, i.e. baseline, associations. See below for a full description of both methods.

#### Brain PC: Change based, memory-sensitive measure

This measure was derived from a sample largely overlapping with that used for the statistical analyses and the AIBL in the present work but included participants down to age > 18 years. Brain PC is based on a principal component (PC) of longitudinal change in 20 cortical thickness and 9 subcortical volume regions. Brain regions were harmonized using a normative modelling framework ^108,109^ with the *PCNtoolkit* (0.30.post2), in *Python3* environment ^110^ (version 3.9.5). This framework offers several advantages as i) it is run independently across sites, ii) can isolate site-effects from other sources of variance associated with it, and iii) produces site-agnostic deviation scores (*z*-statistics) adjusted for age, and sex. *PCNtoolkit* uses a Hierarchical Bayesian Regression (HBR) technique ^111^ and pretrained models from 82 different datasets, including UKB and CamCan data. To avoid losing longitudinal observations, we performed this step recursively by iteratively (n = 100) holding out a calibrating sample and computing the estimates on the remaining data. The average scores of all iterations were used as the standardized scores for each observation. Scanners contributing with < 12 unique individuals or < 25 observations were excluded. For scanners contributing > 12 and < 32 unique individuals, we used a calibration sample consisting of all but 2 participants and estimate the harmonized scores in these two. For scanners with >= 32 unique individuals, we used, in each iteration, a held-out sample of 30 individuals while estimates were applied on the rest.

Next, we selected individuals with at least 2 observations and a minimum follow-up of 1.5 years. For both MRI and memory preprocessed data, we estimated yearly change for each subject, by regressing data on follow-up time. Change data was then fed into separate linear mixed models as implemented in *lme4, lmerTest* ^112,113^, one per brain region. Note that here we used estimates of change, and there was only one observation per individual. For each region, we predicted memory change by brain change, using dataset as random intercepts. Additionally, we used weights to account for potential heteroskedasticity. That is, individuals with short follow-up periods and less observations contribute with more unreliable, high-variance data and thus should produce an unequal spread of residuals. We used the square of reliability as weights as estimated in ^114^. Longitudinal reliability is a function of variance in change and mean measurement error for a given region, and number of observations and total follow-up time for a given individual. After False Discovery Rate (FDR)-correction (p < .05), 29 regions showed significant associations between brain change and memory change, including 9 volumetric subcortical (bilateral amygdala, hippocampus, and thalamus, left lateral and inferior lateral ventricle, right accumbens area) and 20 cortical thickness regions (left G cingul-Post-dorsal, G cingul-Post-ventral, G insular_short, G oc-temp_med-Parahip, G front_inf-Opercular, G front_inf-Triangul, G subcallosal, S temporal_sup; right G Ins lg&S cent_ins, S circular_insula_ant, S oc-temp_med&Lingual, S suborbital; bilateral G temp_sup-Plan_polar, S orbital-H_Shaped, S front_middle, S circular_insula_inf). These regions were entered into the PCA to extract the PC of the memory-sensitive brain regions, yielding a brain measure sensitive to episodic memory change in aging. All regions except the ventricles showed positive loadings with the brain PC.

#### Brain LASSO: Cross-sectional-based, memory-sensitive measure

The alternative brain measure was derived by predicting cross-sectional memory function by cross-sectional brain structure features on an independent sample of UKB individuals not included in other brain analyses. Prediction was performed with a Least Absolute Shrinkage and Selection Operator (LASSO) machine learning algorithm as implemented in the *glmnet* package ^115^. LASSO is a regression technique that performs variable selection and regularization by adding a penalty term, reducing overfitting, and simplifying the model. Lambda was selected as the maximum value within one standard error from minimum lambda, using a cross-validated approach with K = 10 folds (λ = .0143; MSE = .943). LASSO coefficients are provided in the table below. The sample consisted of 28,114 individuals from UKB aged 65.05 years (SD = 7.60) (range 47.32 – 82.78), without longitudinal MRI data, and not included in the main brain analyses. Age was not regressed out allowing prediction to capture both offset and level effects of brain structure on memory function as well as indirect effects due to the unaccounted correlation of age with both MRI features and memory function. We used the Paired associate learning (PAL) (#20197 UKB field) at the first MRI timepoint as index of memory function. MRI data included 337 features; subcortical regions and global brain measures from the *aseg* atlas and cortical area, and thickness regions from the *Destrieux* atlas. Both brain and memory indices were z-standardized, and outliers were considered as values >5 SD apart from the mean. Individuals with outlier values for memory were excluded while brain outlier values were recoded as 0.

**Online table:** LASSO coefficients. Coefficients for the brain measure sensitive to memory derived with a LASSO algorithm using cross-sectional UKB brain structural data predicting memory function as indexed by paired associate learning (PAL; #20197 UKB field). See https://biobank.ndph.ox.ac.uk/showcase/refer.cgi?id=2561 for more information on PAL.

| Region | $\beta$ ($\times 10^3$) |
| --- | --- |
| lh_G&S_frontomargin_thickness | -4.20 |
| lh_G_Ins_lg&S_cent_ins_thickness | 9.71 |
| lh_G_insular_short_thickness | 7.76 |
| lh_G_oc-temp_med-Parahip_thickness | 4.18 |
| lh_G_postcentral_thickness | 3.60 |
| lh_G_precentral_thickness | 21.02 |
| lh_G_temp_sup-Plan_tempo_thickness | 1.04 |
| lh_Pole_occipital_thickness | -44.97 |
| lh_Pole_temporal_thickness | 13.12 |
| lh_S_circular_insula_ant_thickness | 15.11 |
| lh_S_interm_prim-Jensen_thickness | 0.04 |
| lh_S_oc-temp_lat_thickness | -2.01 |
| lh_S_oc-temp_med&Lingual_thickness | 10.74 |
| lh_S_orbital_med-olfact_thickness | -10.29 |
| lh_S_postcentral_thickness | 27.20 |
| lh_S_precentral-sup-part_thickness | 8.70 |
| lh_S_suborbital_thickness | -1.32 |
| rh_G&S_cingul-Ant_thickness | -7.13 |
| rh_G_oc-temp_med-Lingual_thickness | -4.10 |
| rh_G_postcentral_thickness | 8.74 |
| rh_G_temp_sup-Lateral_thickness | 7.73 |
| rh_Pole_occipital_thickness | -9.24 |
| rh_S_circular_insula_sup_thickness | -12.83 |
| rh_S_oc-temp_med&Lingual_thickness | 17.41 |
| rh_S_orbital_med-olfact_thickness | -6.19 |
| rh_S_postcentral_thickness | 16.23 |
| rh_S_temporal_sup_thickness | 0.04 |
| lh_G_and_S_transv_frontopol_area | 7.91 |
| lh_G_cingul-Post-ventral_area | -2.15 |
| lh_G_occipital_middle_area | 7.85 |
| lh_G_temp_sup-G_T_transv_area | 4.10 |
| lh_S_central_area | -1.61 |
| rh_G_and_S_subcentral_area | -1.89 |
| rh_G_subcallosal_area | -13.09 |
| rh_S_central_area | -5.28 |
| rh_S_occipital_ant_area | 4.50 |
| rh_S_orbital-H_Shaped_area | 1.08 |
| rh_S_precentral-sup-part_area | -3.99 |
| rh_S_temporal_sup_area | 4.95 |
| Left-Inf-Lat-Vent | -16.18 |
| Left-Cerebellum-White-Matter | 37.93 |
| Left-Cerebellum-Cortex | 11.86 |
| 3rd-Ventricle | -68.60 |
| Brain-Stem | -23.74 |
| Left-Hippocampus | 17.57 |
| Left-vessel | -2.40 |
| Right-Inf-Lat-Vent | -31.91 |
| Right-choroid-plexus | -27.22 |

### Statistical analyses: Brain cohorts

All the analyses were performed using the R environment (version 4.2.1) ^73^. Visualizations were made with *ggplot2* ^116^ and *ggseg* ^117^ R-packages. Memory, brain variables, and estimated intracranial volume (eTiv) were Z-standardized before inclusion in the models. Outlier values defined as values >5 SD from the mean, were removed from the analyses. Analyses were run using *gamm* models as implemented in the *gamm4 R-package* ^40^, unless otherwise specified.

Memory score was modeled as a function of education, time since baseline, sex, and a dummy regressor for test-retest effects as fixed effects. Baseline age by sex was included as a smooth term. Random intercepts were modeled per participant and dataset, with random slopes of retest effects and time from baseline at a dataset level. To test the effects on memory change, the model was re-run with an additional education × time interaction term. Education was operationalized either as mean-split or based on tertiary education in separate models.

Brain structure was modeled as a function of education, time since baseline, sex, and eTiv as fixed effects. Baseline age by sex was included as a smooth term. Random intercepts were modeled per participant, scanner, and dataset with random slopes of time included at a dataset level. To test effects on brain change, the model was re-run with an additional education × time interaction term. As control analyses, we reran the *gamm* models without eTiv as covariate. Additionally, we ran a linear mixed model as implemented in *lme4,* with eTiv being modeled as a function of education, sex, and baseline age as fixed effects, while site and dataset were included as random intercepts. Only the first observation of each participant was included, as eTiv and education are time-invariant variables. Alternative operationalizations of education and brain structure were tested in separate, but otherwise identical, models.

We used a fuzzy join algorithm, as implemented in *fuzzyjoin* ^118^ to link pairwise MRI and cognitive observations as these were not necessarily collected on the same day. MRI observations were matched with the closest cognitive observations within a maximum time gap of 1 year. Unlinked observations were excluded from the analyses. The relationship between brain, memory level, and education was assessed with several models. *Brain level and memory level:* Memory was modeled by brain structure, sex, time, eTiv, and a dummy regressor for test-retest effects as fixed effects.

Baseline age by sex was introduced as a smooth term. Random intercepts were modeled per participant, scanner, and dataset with random slopes of retest and time modeled at a dataset level. *Brain change and memory change:* An additional brain × time term was added to the model. *Moderating effect of education on level – level associations:* Additional terms for education and education × brain were added in the first model. *Moderating effect of education on change – change associations:* A triple interaction term (brain × time × education) as well as its lower order components were added in the first model. *Control analyses:* A main education term – without any interaction – was added to the models to assess level – level and change – change associations between brain and memory, to test whether the strength of these associations was affected by education level. As with other analyses, alternative operationalizations of education and memory-sensitive brain structure were tested in separate but comparable models.

